# Prediction of Covid-19 Infections Through December 2020 for 10 US States Using a Two Parameter Transmission Model Incorporating Outdoor Temperature and School Re-Opening Effects

**DOI:** 10.1101/2020.08.06.20169896

**Authors:** Ty Newell

## Abstract

Covid-19 infection case predictions (total cases) are made for August through December 2020 for 10 US States (NY, WA, GA, IL, MN, FL, OH, MI, CA, and NC). A two-parameter model based on social distance index (SDI) and disease transmission efficiency (G) parameters is used to characterize SARS-CoV-2 disease spread. Current lack of coherent and coordinated US policy causes the US to follow a linear infection growth path with a limit cycle behavior that modulates the US between accelerating and decaying infection growth on either side of a linear growth path boundary.

Four prediction cases are presented:

1. No school re-openings; fall season temperature effect
2. No school re-openings; no fall season temperature effect
3. School re-openings; fall season temperature effect
4. School re-openings; no fall season temperature effect

Fall outdoor temperatures, in contrast to the 1918 pandemic, are predicted to be beneficial for dampening SARS-CoV-2 transmission in States as they pass through “swing season” temperature range of 70F to 50F. Physical re-opening of schools in September are predicted to accelerate infections.

States with low current infectious case numbers (eg, NY) are predicted to be minimally impacted while States with high current infectious case numbers (eg, CA and FL) will be significantly impacted by school re-openings.

Updated infection predictions will be posted monthly (Sept, Oct, Nov, Dec) with adjustments based on actual trends in SDI and G. Assessments related to outdoor temperature impact, school re-openings, and other public gathering re-openings will be discussed in updated reports.

## Introduction

The goal of this paper is projection of potential Covid-19 infection cases from August through December 2020 in 10 US States (NY, WA, GA, IL, MN, FL, OH, MI, CA, NC) that represent a variety of infection case histories and trajectories. A two-parameter prediction model has been developed to provide a conceptual framework for understanding two primary factors that affect the spread of SARS-CoV-2. A Social Distance Index (SDI) and a disease transmission efficiency (G) are the two parameters that define the growth and decay of SARS-CoV-2 infection.

Accurate prediction of the spread of SARS-CoV-2 can be made based on the Social Distance Index (SDI) and a virus transmission efficiency parameter (G). The two parameters define an Infection Parameter, IP, that is analogous to the basic Reproduction Number, Ro. The Infection Parameter has exposure time (1 week assumed) and infectious period (2 weeks assumed) built into its formulation. An IP value of 2.72 (“e”) results in linear total infection growth that is a mathematical boundary between increasing and decreasing new infection cases per day. The mathematical formulation and correlation procedures for relating disease infection dynamics to SDI and G are described in a previous MedRxiv posting (1).

Sustained IP values below 2.72 result in continuous decay of infectious cases. Lower IP levels decrease new infections at an increased rate. An IP value of 1 represents either perfect isolation of susceptible subjects from infectious subjects or zero transmission efficiency of the virus, leading to total decay of infectious cases. Several countries with coherent, coordinated disease management programs (strict isolation, wearing of face masks, etc) have successfully maintained IP levels below 2.72, resulting in infection case reductions to levels in which testing, tracing and isolation programs can be established for continued control of virus infections.

The lack of a coordinated, coherent nationwide program for Covid-19 disease management in the US has resulted in linear daily growth of total infection cases. Figure 1 shows US total infections relative to total infections in eight other countries that have been able to maintain IP levels below 2.72 for sustained periods.

**Figure 1.**
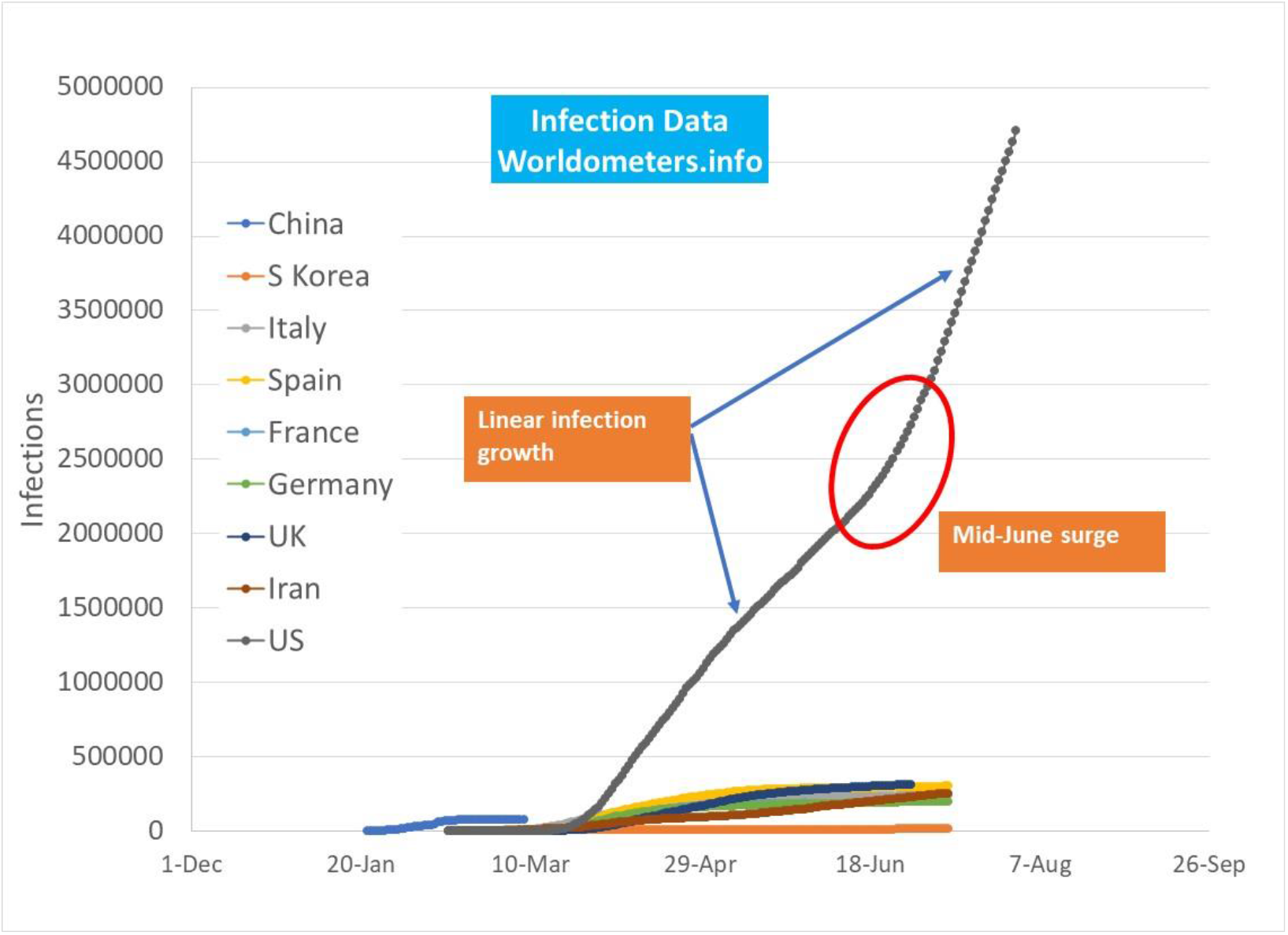
US total infections as of July 31 2020 with linear infection path regions before and after mid-June “summer surge”.

Two linear infection growth regions for the US are shown in Figure 1. The earlier linear region developed as accelerated infection growth in New York, Illinois, Michigan and other early impacted states were able to reduce infection growth rates. The second linear infection growth region developed after an accelerated surge of infections occurred in mid-June due to a combination of increasing outdoor temperatures (1), overly aggressive re-opening of public gathering places, and lax personal protection measures (face masks).

Regionally, US States appear to be operating in a limit cycle that alternately crosses the linear boundary between accelerating and decelerating infection growth rates. Opposing forces pushing and pulling a region across the infection growth/decay boundary are the pressure to re-open public gathering spaces (schools, businesses, religious meeting places, etc) and the public’s reaction to uncontrolled, accelerated infection growth rates.

Accelerated US infection growth occurred in mid-June due to pre-mature business openings in some States and an outdoor temperature effect related to people moving indoors as outdoor temperatures increased above 70F. The “summer surge” moved the US to the second linear infection growth path shown in Figure 1. Note that any linear infection growth path, regardless of slope, is a boundary between accelerating and decelerating (decay) daily total infections. That is, when a transient effect occurs and daily infection growth becomes non-linear, the populace of that region tends to re-establish linear daily infection growth at some other slope.

The upcoming fall and winter seasons in the US are anticipated to disrupt infections throughout the country as outdoor temperatures change and pressure to re-open schools and businesses builds. Based on the US current affinity for operating along a linear infection boundary path, infection predictions for August through December 2020 are presented for four cases:

1. No school re-opening; outdoor temperature effect
2. No school re-opening; no outdoor temperature effect
3. School re-opening; outdoor temperature effect
4. School re-opening; no outdoor temperature effect

During the 1918 pandemic, a deadly fall infection “wave” occurred. In contrast to the 1918 pandemic, the current pandemic’s autumn outdoor temperature effect is predicted to create a beneficial, temporary drop in disease transmission efficiency. As outdoor temperatures decrease below 70F (21C), windows will be opened and fresh air ventilation will increase, minimizing indoor contagion concentration. Also, people will spend more time outside where disease transmission efficiency is reduced. Note that people spending time outside has the beneficial effect of reduced indoor contagion release. Homes and buildings will be re-closed and sealed as outdoor temperatures decrease below 50F, causing an increase in disease transmission efficiency that increases infection growth.

The second effect examined with the prediction model is the anticipated physical re-opening of schools. At present, it is unknown how many schools will re-open, as well as how physical interactions of people will be controlled. The two-parameter prediction model allows estimation of school re-opening impacts through an adjustment to the Social Distance Index (SDI). We apply a fixed, 20% reduction of the SDI to account for increased social interaction during the month of September. The assumed increase of social interaction represents a level that is intermediate between today’s SDI level, and SDI levels that existed prior to Covid-19 in February 2020. At present, much uncertainty revolves around physical re-opening of schools and businesses. The current prediction provides a reference that will be updated monthly as trends in the SDI parameter become known.

## Infection Parameter (IP), Social Distance Index (SDI) and Disease Transmission Efficiency (G)

A complete discussion of the prediction model’s formulation, parameter correlation methods, and sources of data are discussed in reference (1). A qualitative description of the model and parameter relations is provided here to explain characteristics of the predictive model relative to outdoor temperature and school re-opening effects.

All data used for modeling and correlations are publicly available. Data for US total infections are obtained from Worldometers.com that compiles data from multiple sources (2). Data for US States total infections are obtained from 91-divoc.com, a website formulated by a University of Illinois faculty member that utilizes data from Johns Hopkins University (3). Data for the Social Distance Index is obtained from the University of Maryland’s Transportation Institute website (4).

Figure 2 shows the trend of the US Infection Parameter (IP) since March 2020. Conceptually, IP is the ratio of new infections over a 14 day period per infectious person. Initially, IP was very high prior to social isolation and implementation of disease transmission control measures. Also, unidentified asymptomatic cases and inadequate testing to find infections result in relatively high IP values. For example, 30 new infections per infectious person during early Covid-19 incursion in the US may actually be 30 new infections relative to 1 known infectious person and 4 unknown infectious persons. Current IP values may still represent “hidden” infectors, however, these effects do not detract from the prediction model’s ability to accurately project future infections.

**Figure 2.**
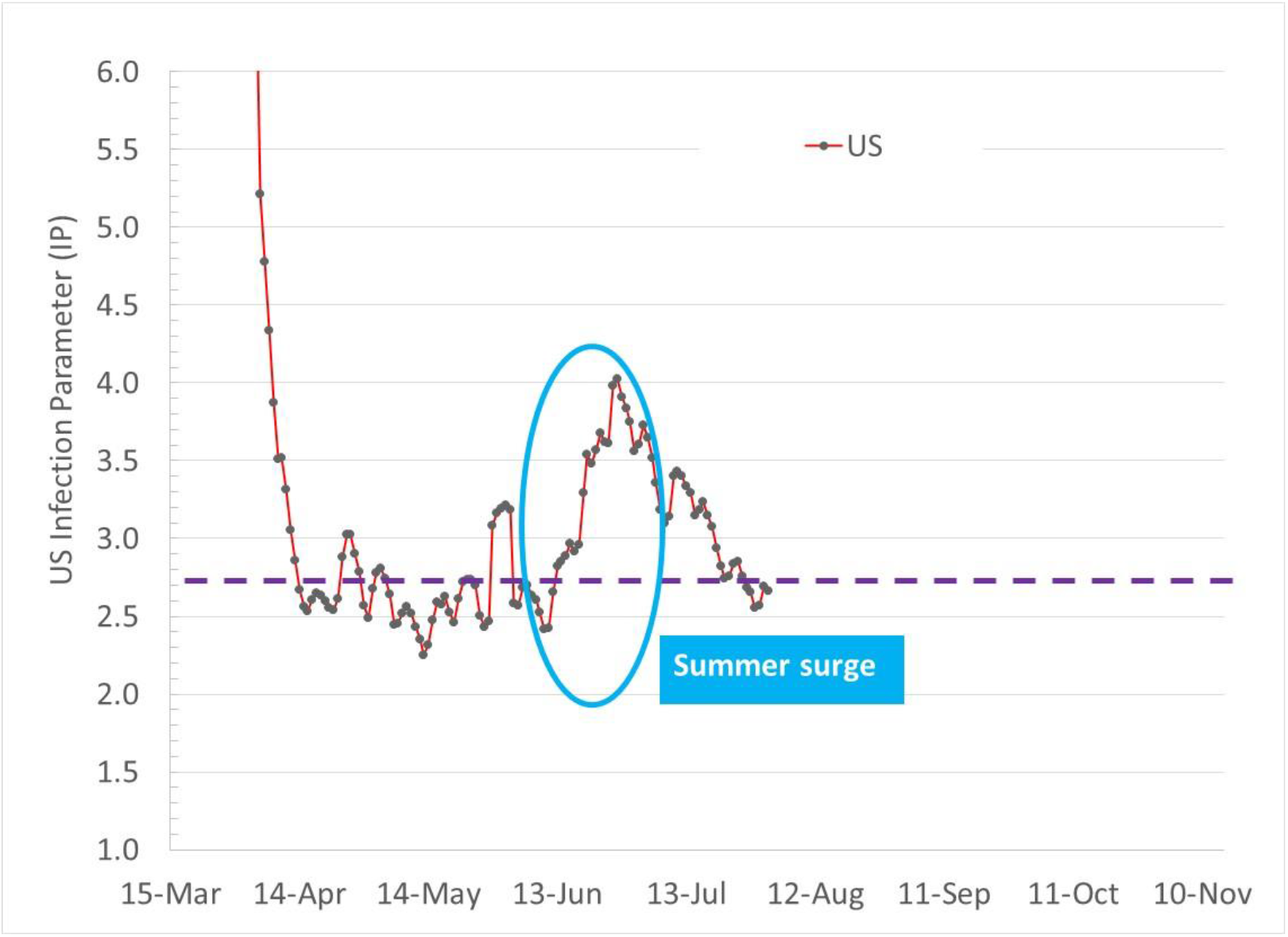
US Infection Parameter (IP) showing tendency to move along linear total infection growth boundary (IP=2.72). The mid-June “summer surge” caused by a combination of outdoor temperature increase above 70F and pre-mature business openings is followed by local populace actions to reduce infection growth back to the linear growth boundary.

Figures 3 and 4 show IP trends for some States since March 2020. Figure 3 shows IP trends for 3 States (CA, FL, and NC) that have struggled to reduce IP levels below the linear IP boundary of 2.72. Florida, with aggressive re-openings and lax interpersonal disease transmission control measures began experiencing rapid infection growth in early June. A long incubation period followed by a 2 week infectious period delays a locale’s realization of an accelerated infection trajectory. Once a region identifies accelerated infection growth, control measures will not produce immediate reductions in infections due to the week long incubation period and 2 week infectious period.

**Figure 3.**
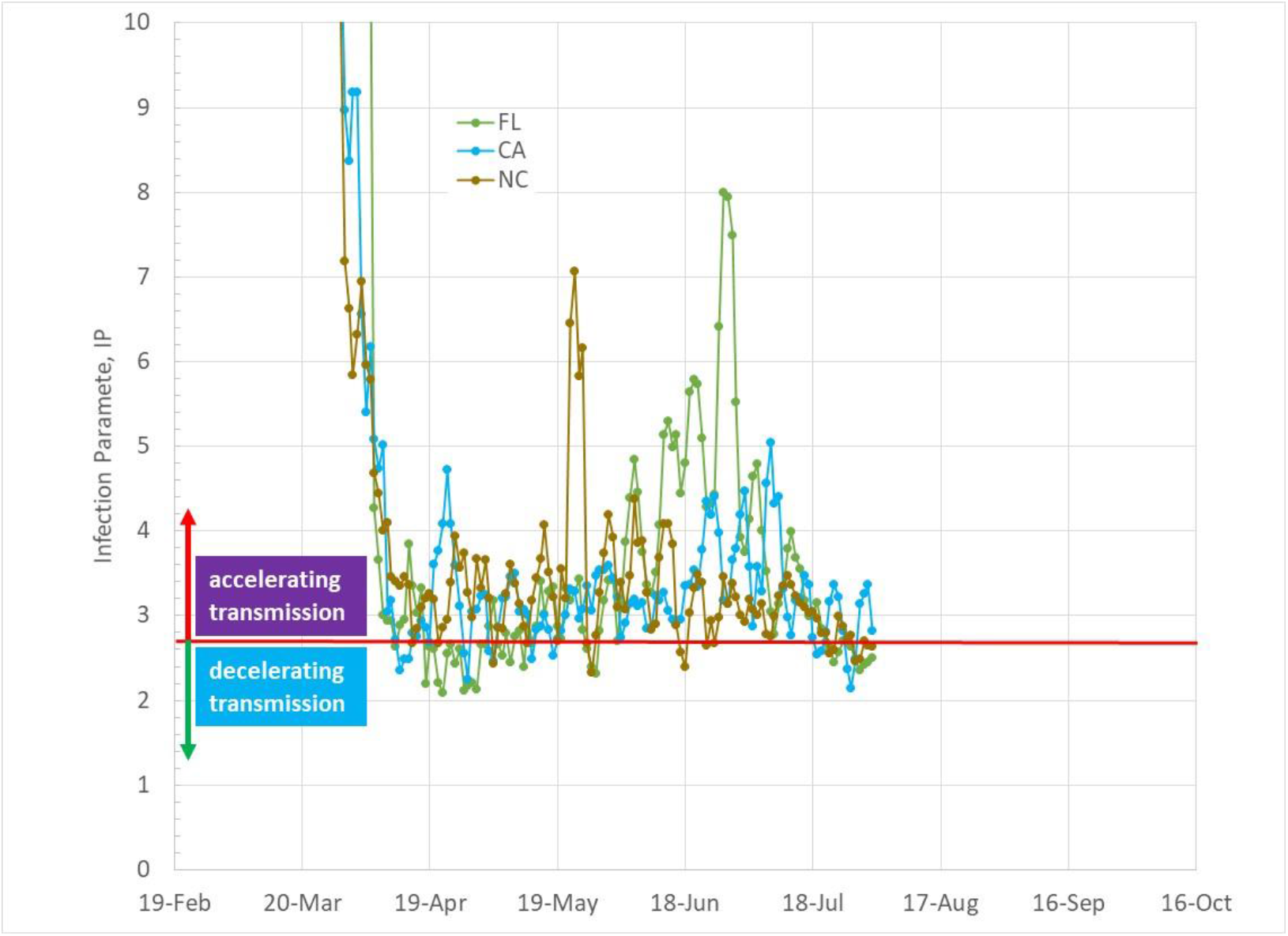
Daily Infection Parameter trends for 3 US states that re-opened, causing IP increases, followed by actions to reduce infection growth, returing to linear infection growth boundary (IP=2.72).

**Figure 4.**
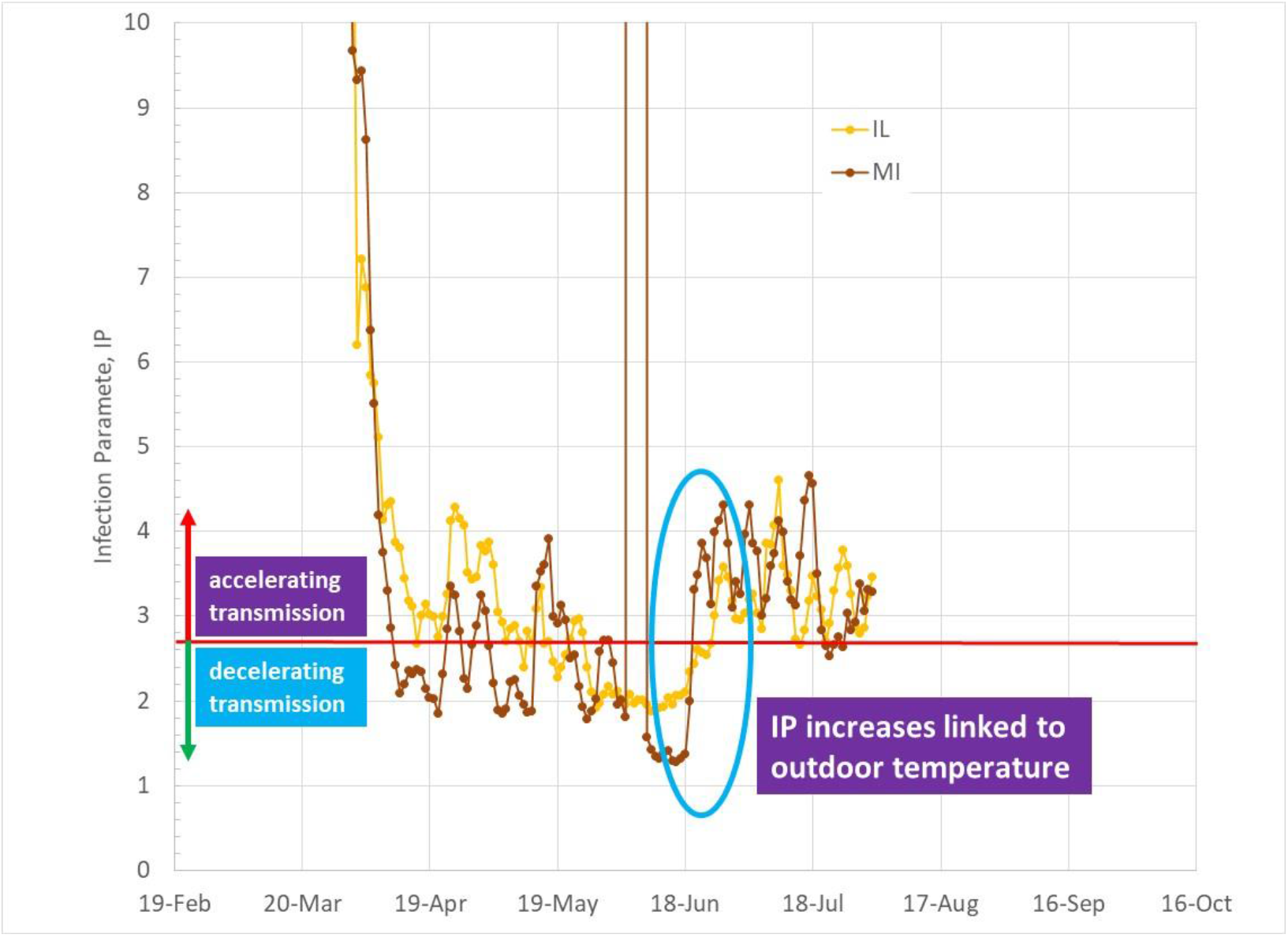
Daily Infection Parameter trends for 2 US states that show strong IP increases as outdoor temperatures increased above 70F (21C). Note that the sharp IP increase in MI during early June is due to a change in data processing.

Figure 4 shows the characteristic jump in IP in States such as Illinois and Michigan related to outdoor temperatures increasing above 70F (21C). Both Michigan and Illinois had reduced IP below 2.72 by the end of May, and were experiencing sustained decay of infections. An unexpected IP jump occurred in both states in mid-June, moving IP back into the accelerated infection growth region. Both Illinois and Michigan struggled to again reduce IP through additional disease transmission mitigation measures and have nearly reached the linear infection growth boundary line as of the end of July.

Figures 5, 6, and 7 show US daily IP values as functions of a Social Distance Index (SDI), a parameter obtained from the University of Maryland’s Transportation Institute (4), and a defined disease transmission efficiency parameter (G). The two parameters provide a conceptual framework for understanding independent, physical actions that impact the Infection Parameter. SDI, derived from anonymous cellular and vehicular gps data, measures the amount of travel and type of trips (eg, travel for work, shopping, socializing) taken by a region’s populace. An SDI of 0 represents maximum social interactions while large SDI values indicate increased isolation.

**Figure 5.**
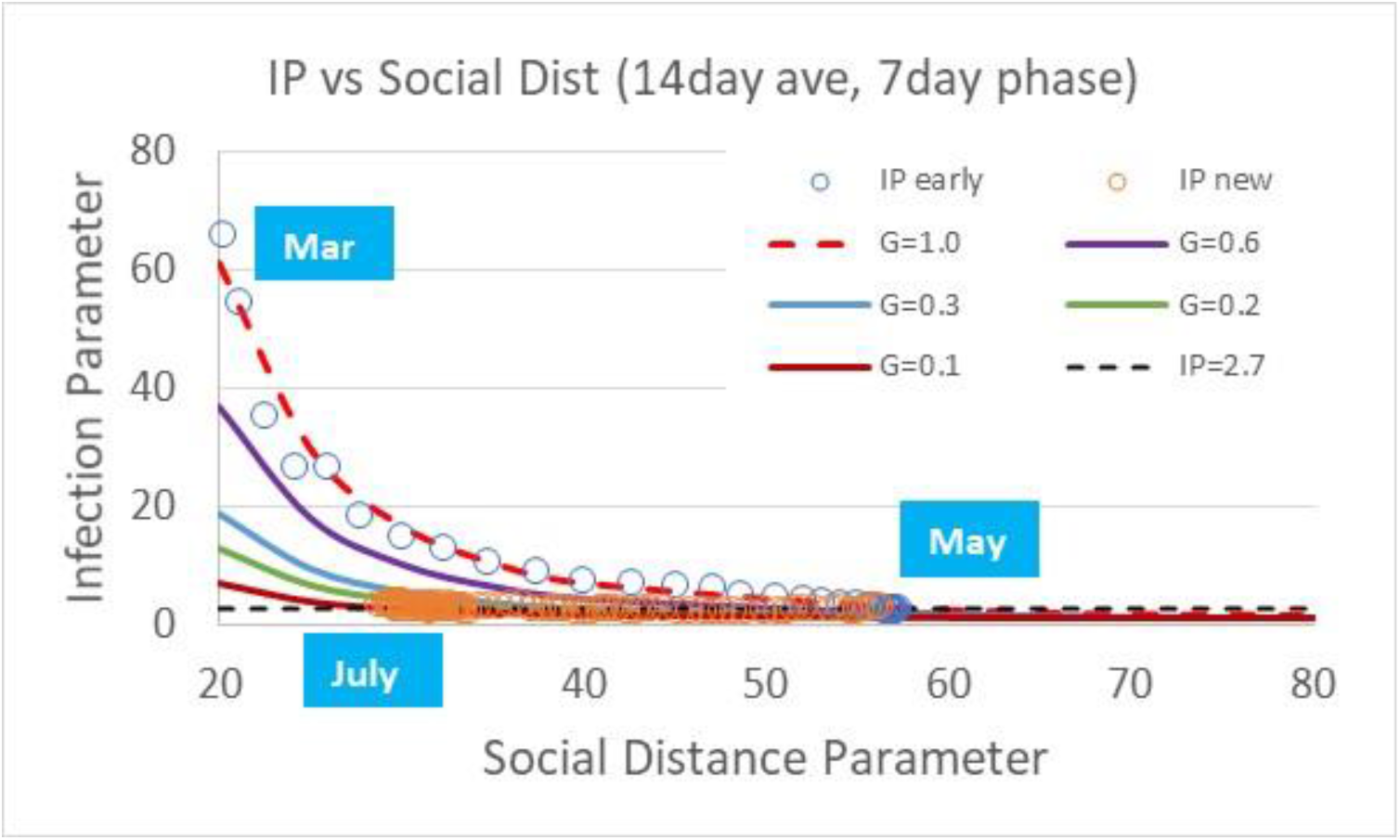
US Infection Parameter trends as a function of the Social Distance Index (SDI)

**Figure 6.**
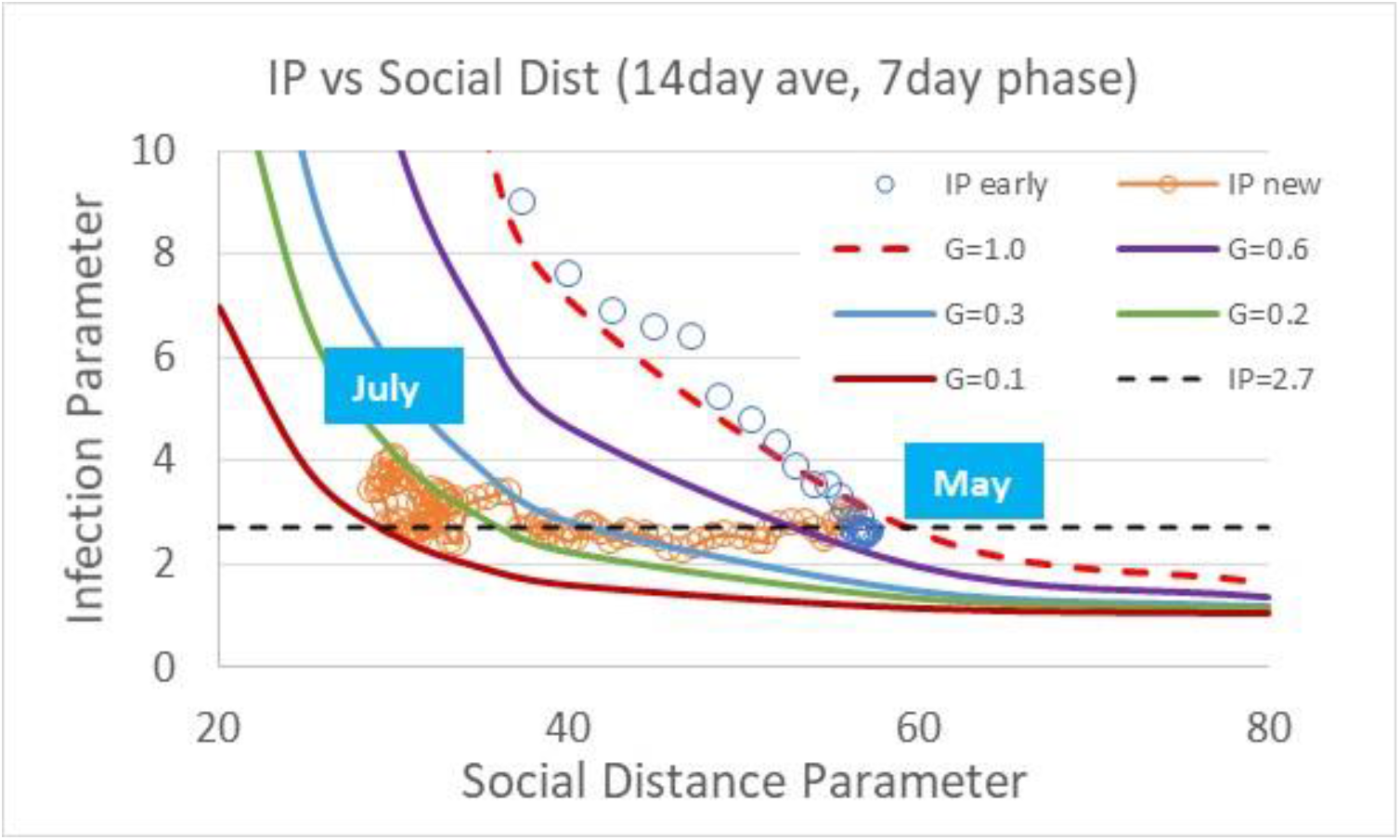
Amplified view of US Infection Parameter trends as a function of Social Distance Index.

**Figure 7.**
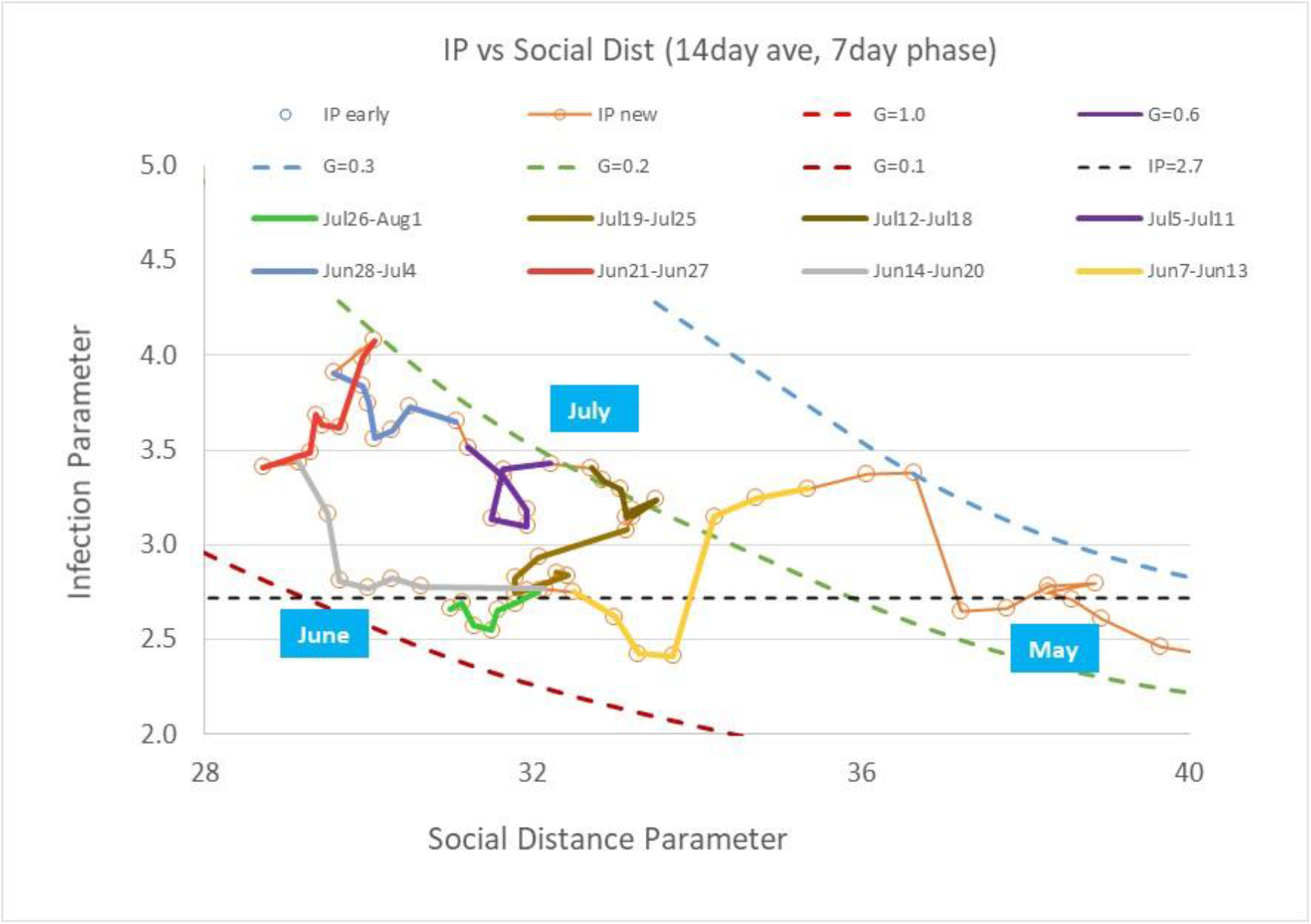
US daily Infection Parameter trends as a function of Social Distance Index from May through July.

Figure 5 shows the daily IP path relative to SDI for the US since March. The initial data (open blue data points) during March and April occurred when much of the US began isolating, but before additional measures such as face masks and 6ft distancing were implemented. The path outlined by the open, blue circle data is used to define a disease transmission efficiency parameter, G, with an assigned value of unity as shown in Figure 5 by the dashed red line. Parametric values for other disease transmission efficiencies are relative to the reference efficiency.

Figure 6 is an amplified view of the US IP path showing the progression as the US went into isolation from March through April, followed by re-opening of the US coupled with implementation of disease transmission control measures. Beginning in May, and continuing through July 2020, SDI decreased, indicating that people were traveling and interacting more. As re-openings occurred, disease control measures such as face masks, reduced indoor occupancy, surface sanitation, 6 ft spacing, and other practices reduced the disease transmission efficiency such that IP remained low.

Figure 6 shows the US tendency to move along an IP value of 2.72, the boundary between accelerated infection growth and decaying infections. Figure 7 focuses on more recent data that covers late May through July in which the first and second linear infection growth paths shown in Figure 1 occurred. As the US re-opened, SDI decreased, with IP alternately moving above and below the 2.72 linear infection growth boundary. By mid-June, SDI had decreased to 29. Over the course of a week, as many states warmed to temperatures above 70F, an increase of disease transmission efficiency occurred without significant change of SDI. The increase of G has been linked to the movement of people into inadequately ventilated, air-conditioned buildings as outdoor temperature elevated above 70F (21C) (1). The increase in IP was sufficient to initiate a new infection surge in states that had successfully reached the infection decay region.

A relation correlating IP with SDI and G has been developed (1):

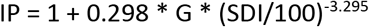

Note that a relation between G and SDI representing the linear infection growth boundary line that divides accelerated infection growth and decaying infections can be obtained by substituting IP=“e”, resulting in:

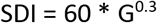

This relation shows, for example, that a disease transmission efficiency of 0.5 requires an SDI of 49, while a disease transmission efficiency of 1 requires an SDI of 60. Note that disease transmission efficiency can be greater than 1, as observed in densely populated regions, such as New York City where mass transit, denser housing, and other factors enhance disease transmission relative to social distancing metrics.

## Linear US Infection Growth and Infection Parameter Limit Cycle

Figure 7 shows the collective IP path of the US in terms of SDI and G parameters from May through July. The outdoor temperature surge in mid-June coupled with a low SDI (SDI~29, more social interaction) from re-openings elevated IP to nearly 4, causing rapid infection growth. Colored path segments show how the US returned to the linear infection growth path of IP=2.72. Initially, as accelerated, uncontrolled infection growth was reported in the news, people reacted by increasing SDI (increased isolation). Late June and early July colored path segments in Figure 7 trace a constant transmission efficiency path with G~0.19 as SDI increased to 33. The associated decrease of IP slowed infection growth.

Renewed pressure to continue allowing businesses to re-open, coupled with efforts to enforce face masks, reduced indoor occupant density, movement of restaurant tables to outside, and other measures resulted in a combined decrease in SDI and decrease in G. As of the end of July, SDI has been reduced to 31 and G has been reduced to 0.12, resulting in an IP somewhat below the linear infection growth boundary of 2.72.

Individual states followed various paths throughout the pandemic’s history in the US, however, most tend to show a similar behavior of oscillating across the linear growth boundary. Figure 8 shows the path taken so far by Michigan. The spurious, off-scale jump in IP is due to a change in data reporting. Michigan’s initial reduction in IP during March and April followed a path above G=1, reflecting the concentration of infections around Detroit, similar to New York City’s elevated disease transmission efficiency. As Michigan gained control, sustaining IP levels below 2.72, significant infection case decay occurred. Note that Michigan also experienced a beneficial drop in IP associated with outdoor temperatures rising above 50F as discussed in reference (1). As Michigan began re-opening businesses from May to mid-June, and outdoor temperatures increased above 70F, a sharp elevation in IP occurred that is linked to people moving into buildings, closing windows and reducing fresh air ventilation. The jump of IP moved Michigan and several other states into an accelerated infection growth region. Alarmed Michigan citizens reacted with a slight increase of SDI coupled with renewed efforts to decrease G (ie, increased face mask wearing).

**Figure 8.**
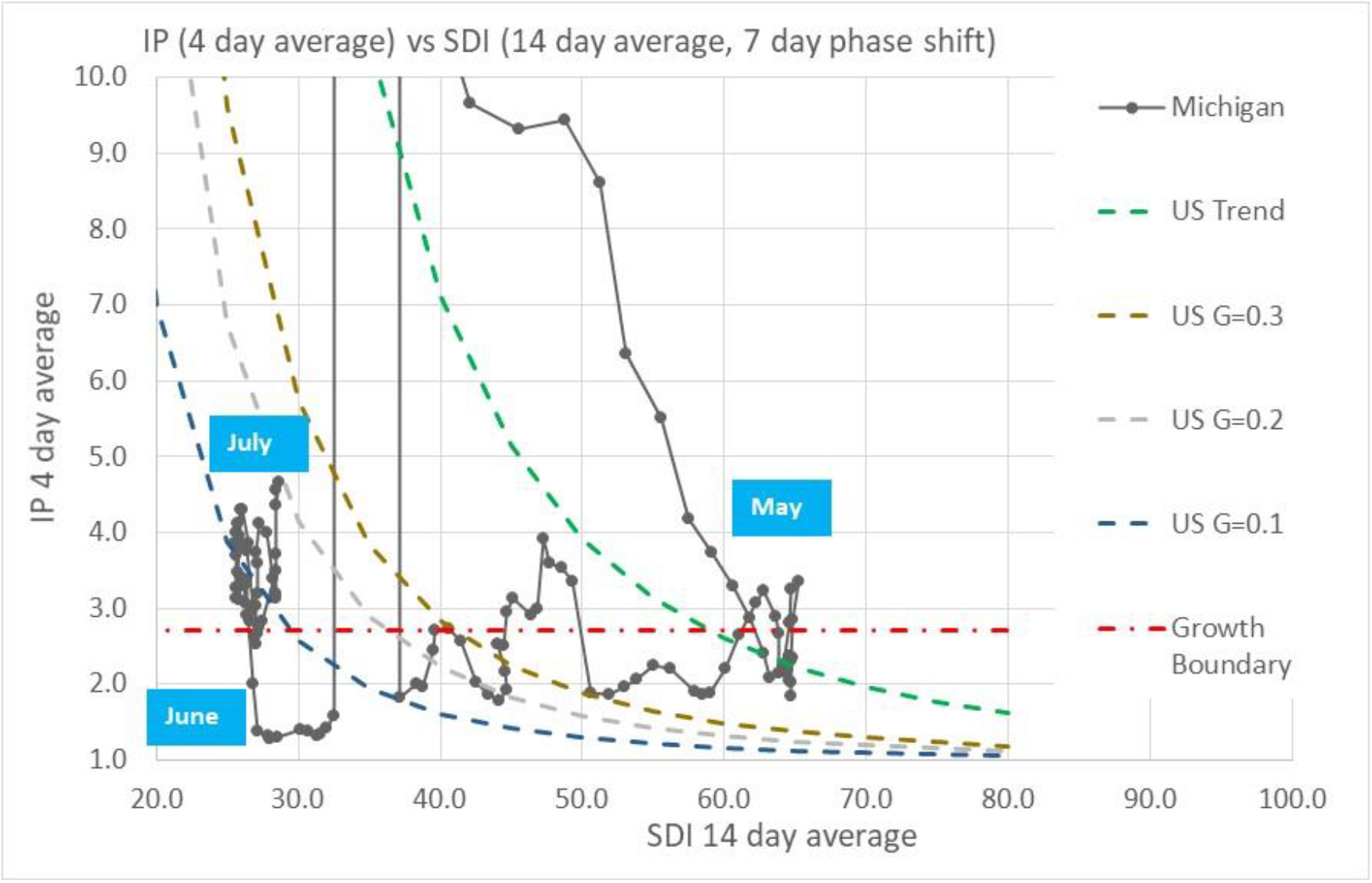
Michigan daily Infection Parameter trends as a function of Social Distance Index.

## Prediction of Total Covid-19 Infections and New Daily Infection Cases Through December 2020

Projection of Covid-19 infection case trends for 10 US States through December 2020 are made based on the following assumptions:

1. US States continue oscillating across the linear growth boundary separating infection accelerated growth and decay regions (IP=2.72)
2. The disease transmission efficiency parameter, G, is reduced by 25 to 33% when outdoor temperatures are in the “swing season” temperature range of 50F (10C) to 70F (21C)
3. Physical re-opening of schools causes a reduction in a State’s SDI of 20% along a line of constant disease transmission efficiency

Based on these three conditions, we project infection trends for the following four cases:

1. No school re-opening; outdoor temperature effect
2. No school re-opening; no outdoor temperature effect
3. School re-opening; outdoor temperature effect
4. School re-opening; no outdoor temperature effect

Figures 9 and 10 show actual SDI values from early March through July 2020 followed by assumed SDI paths from August through December 2020 for 10 US States. Figure 9 shows SDI trends assuming no physical re-opening of schools. Figure 10 shows the assumed 20% reduction of SDI values relative to unadjusted SDI values for the month of September. The 20% reduction is a change of SDI that is higher than “normal” (pre-Covid-19) SDI values in early March. Actual SDI changes caused by physical re-opening of schools and businesses are unknown, and the temporal distribution of the effect is uncertain as many schools districts may roll out a gradual transition from virtual classrooms to physically occupied classrooms. Comparison between the “no school” predictions and “school” predictions provide some perspective of the magnitude of differences between these cases.

**Figure 9.**
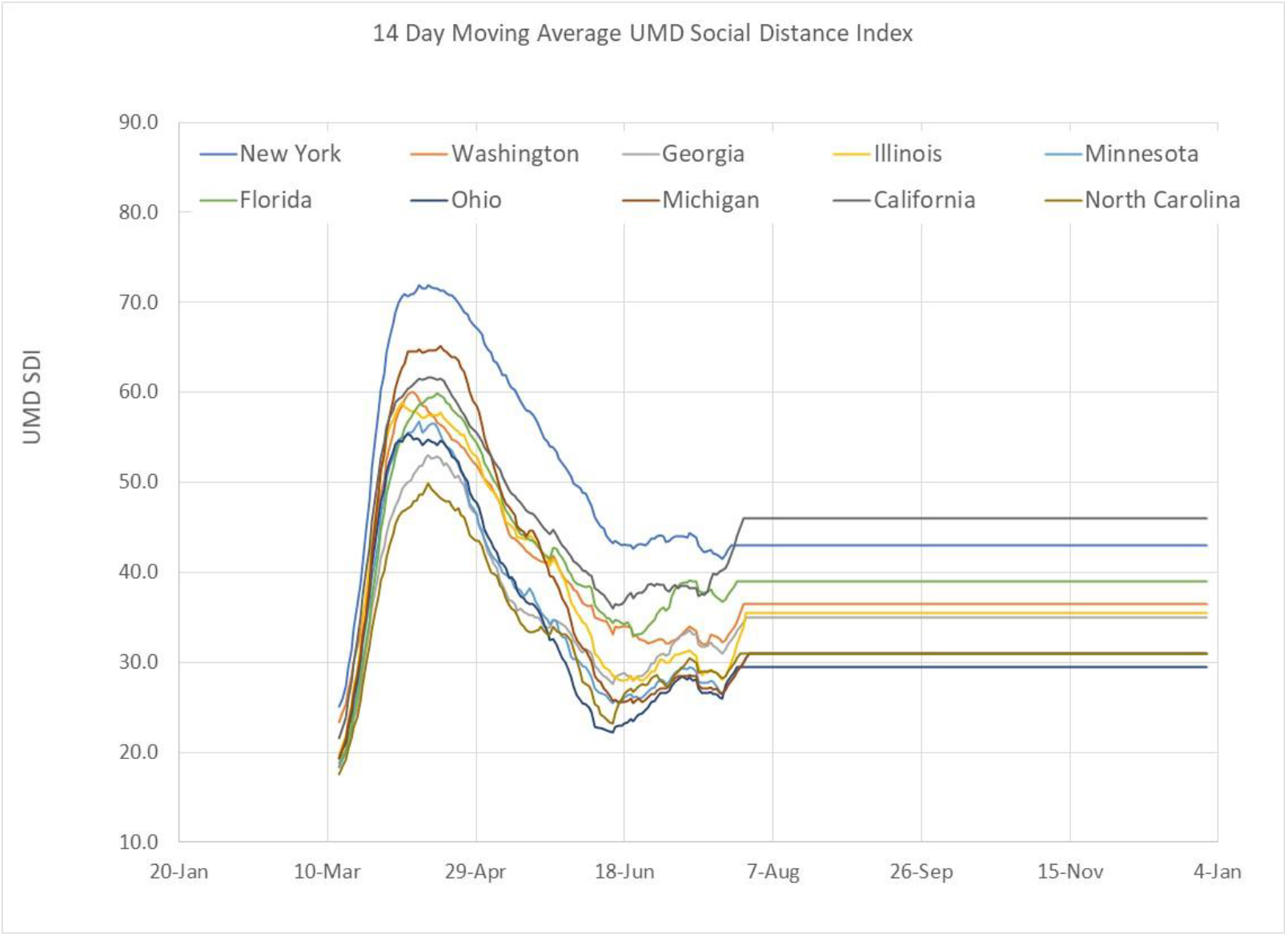
Social Distance Index (SDI) trends for 10 US States projected from August through December 2020 based on SDI levels at the end of July without adjustment of SDI for school re-openings.

**Figure 10.**
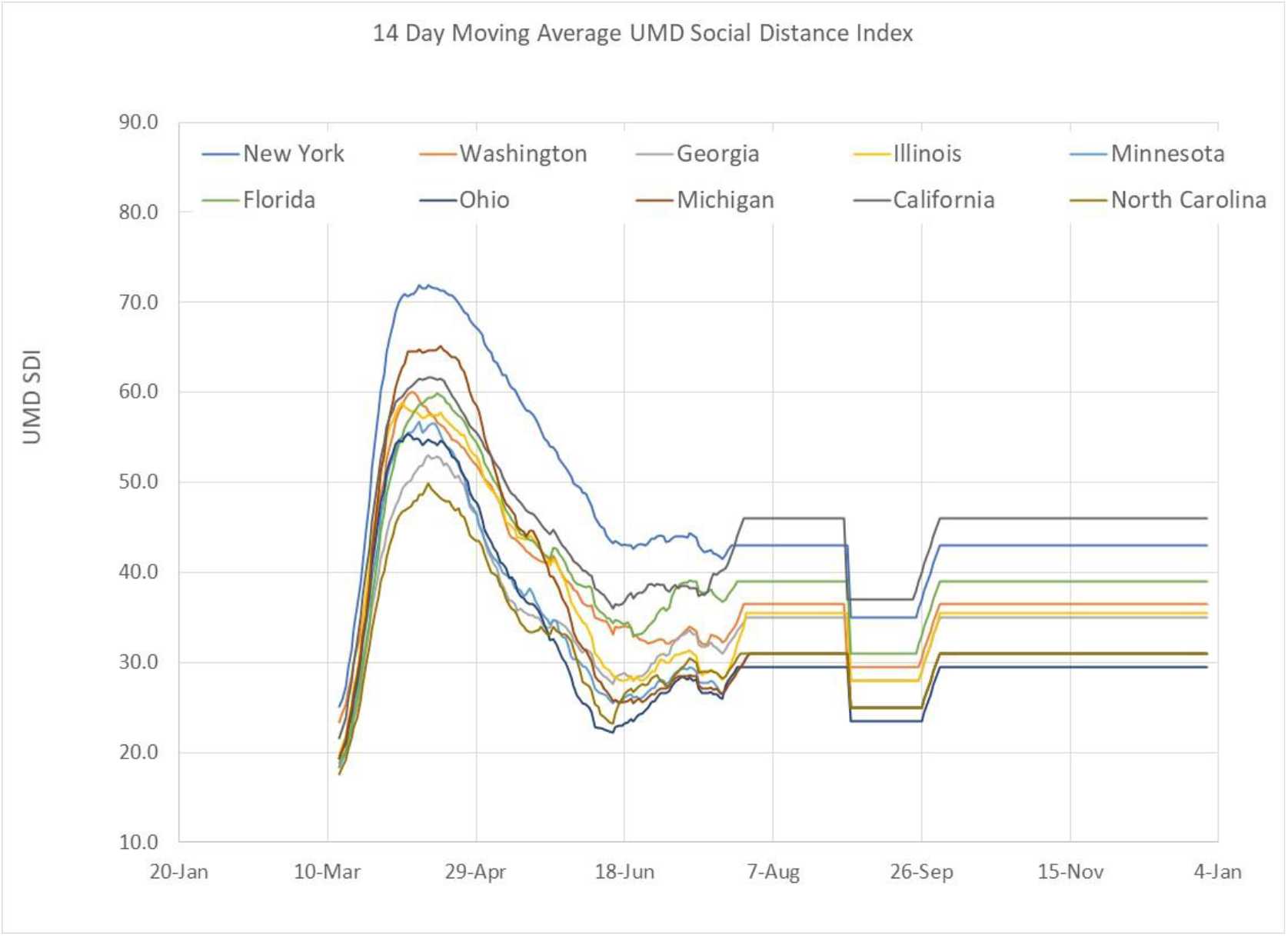
Social Distance Index (SDI) trends for 10 US States showing a 20% reduction of SDI during the month of September representing increased social interactions related to school re-openings.

The disease transmission efficiency, G, has been selected based on current (end of July 2020) levels. In addition, slight adjustment to SDI and G have been made that results in an Infection Parameter below the linear infection growth boundary. In this manner, projected SDI and G levels are held constant along the linear infection growth boundary path, as is the observed current behavior tendency in the US. Outdoor temperature effects of 25 to 33% reduction of G are made for each location relative to when historic weather data indicates that State is between 50F (10C) and 70F (21C). MI, IL, and OH displayed stronger outdoor temperature effects than other states, with G reduced by 33% during the 50F (10C) to 70F (21C) swing season window. All other states had G reduced by 25% during swing season conditions.

Figures 11–18 show Infection Parameter trends for 10 States for combinations of school and no school effects coupled with outdoor temperature and no outdoor temperature impacts. Figures 11 and 12 show “school” and “no school” re-opening IP trends for NY, WA and Ga projected through the end of 2020. Washington is entering the swing season window in August, with New York and Georgia entering the swing season temperature window in September. At the end of October and beginning of November, Washington will drop below 50F and exit the temperature window with IP increasing as citizens closed buildings and stay inside more regularly. New York, followed by Georgia will also exit the temperature window as temperatures lower in approximately 1 to 2 week succession as observed in Figure 11.

**Figure 11.**
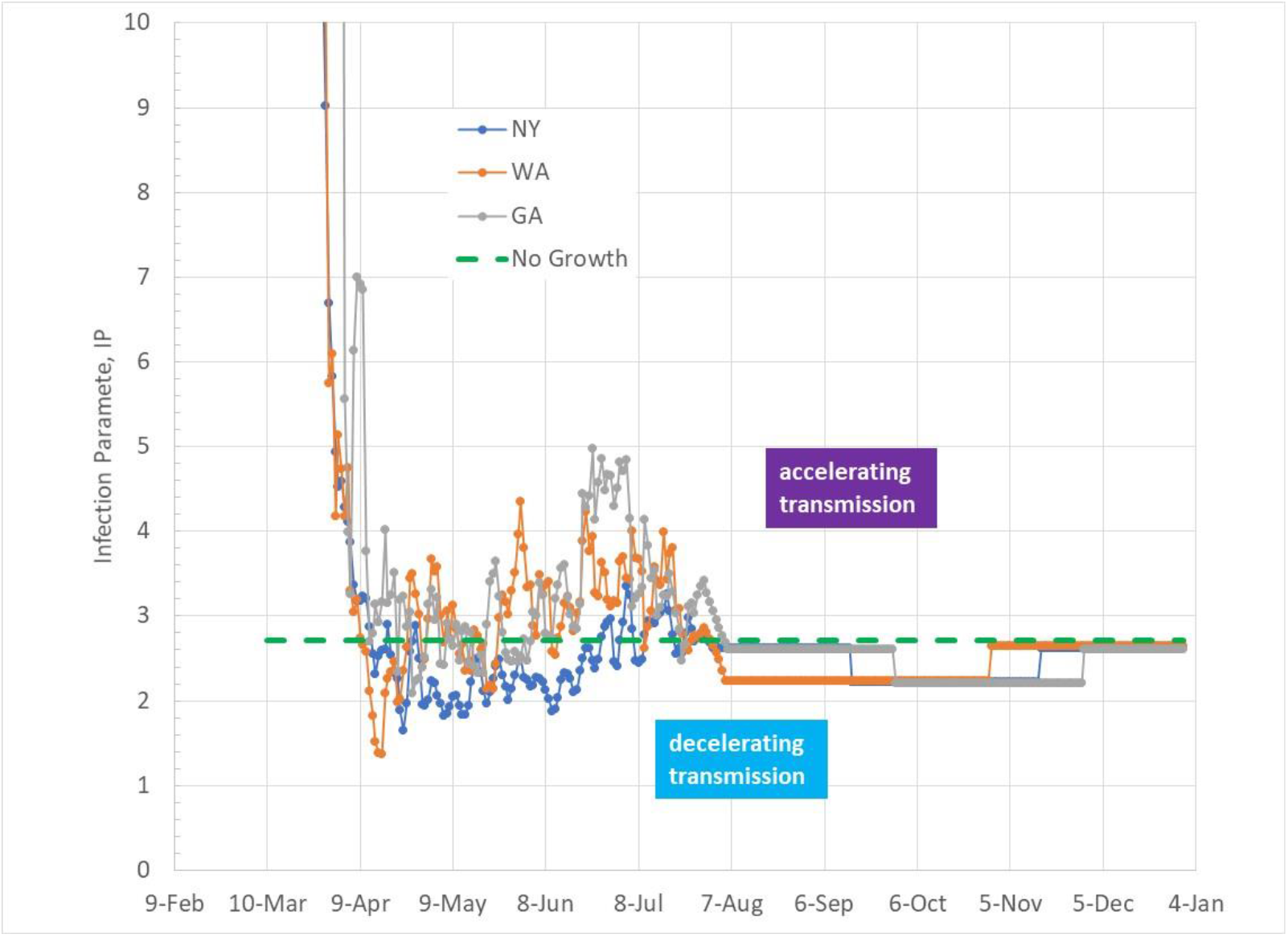
Infection Parameter projections for NY, WA, and GA showing impact of outdoor temperature without school re-opening from August through December 2020.

**Figure 12.**
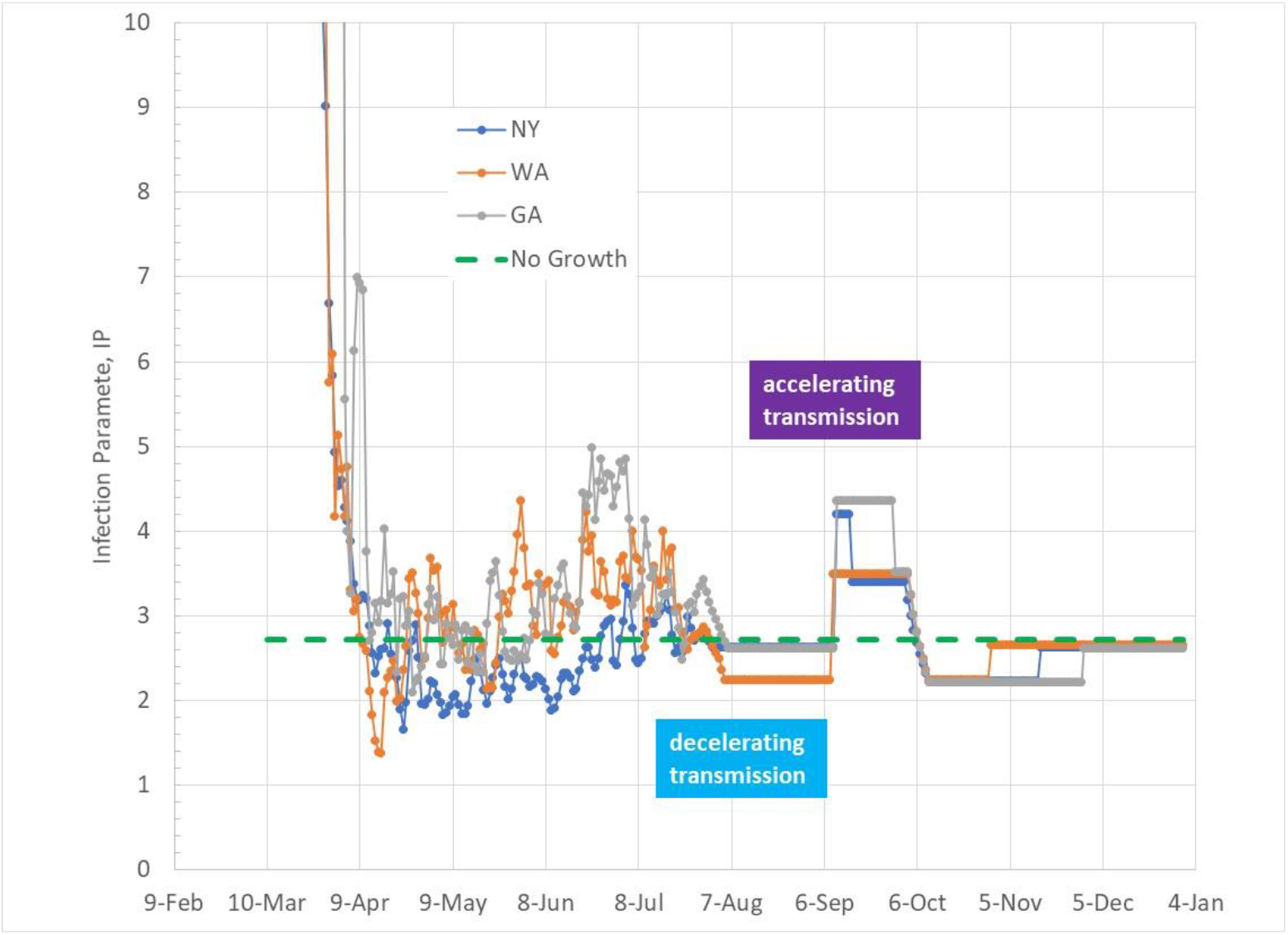
Infection Parameter trends for NY, WA, and GA including the effect of outdoor temperature coupled with a 20% SDI reduction during September.

**Figure 13.**
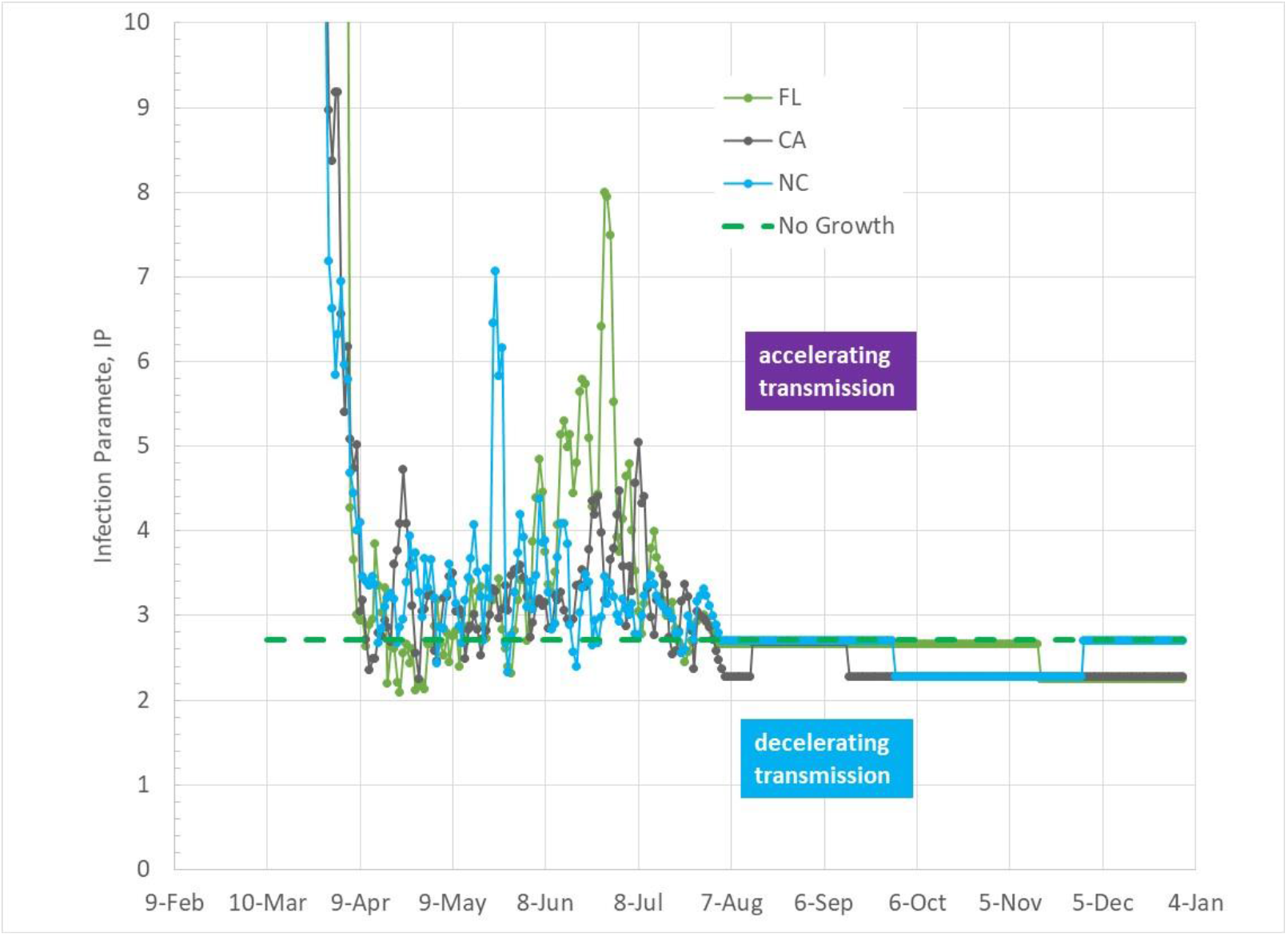
Infection Parameter projections for FL, CA, and NC showing impact of outdoor temperature without school re-opening from August through December 2020.

**Figure 14.**
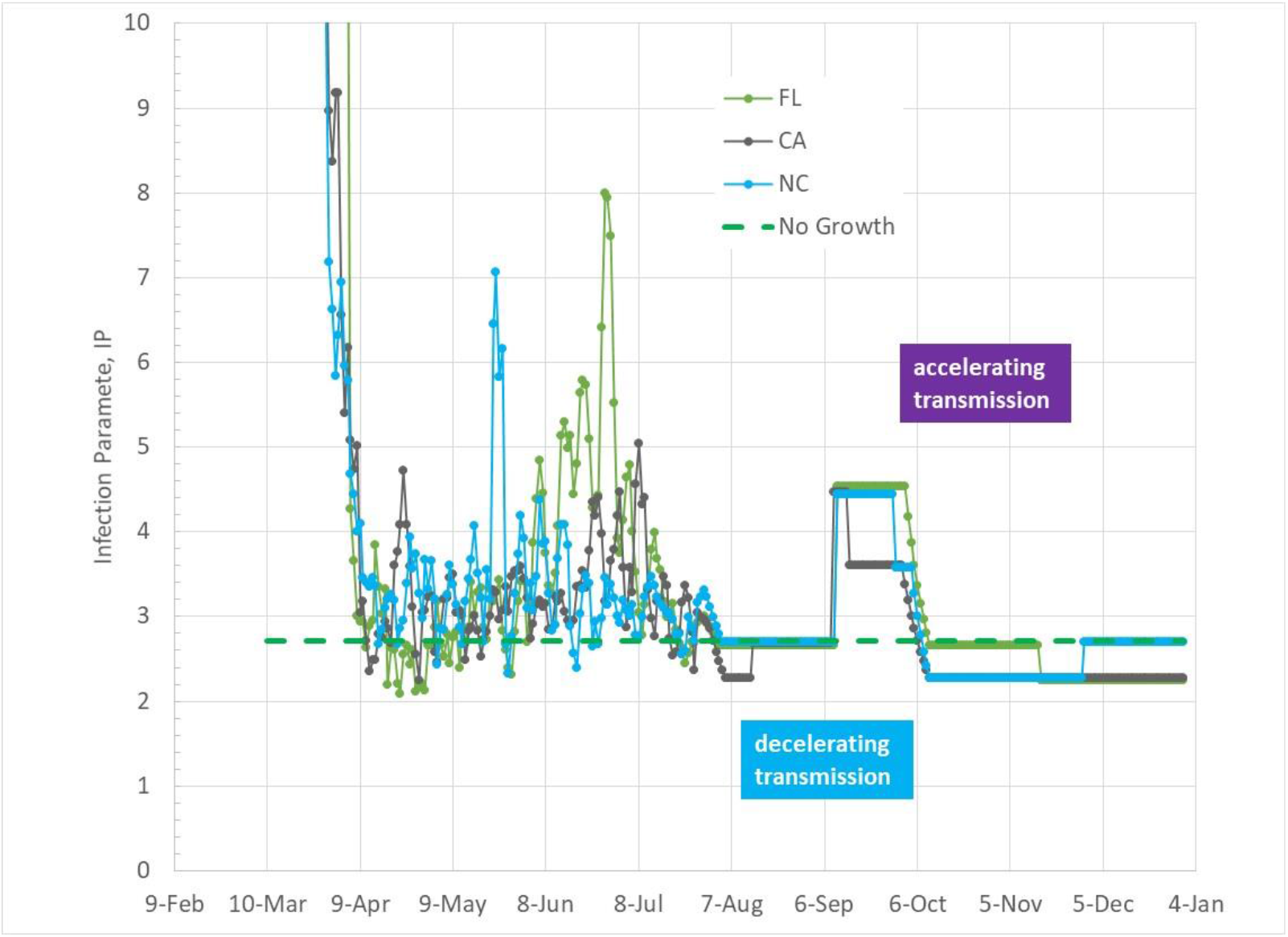
Infection Parameter trends for FL, CA, and NC including the effect of outdoor temperature coupled with a 20% SDI reduction during September.

**Figure 15.**
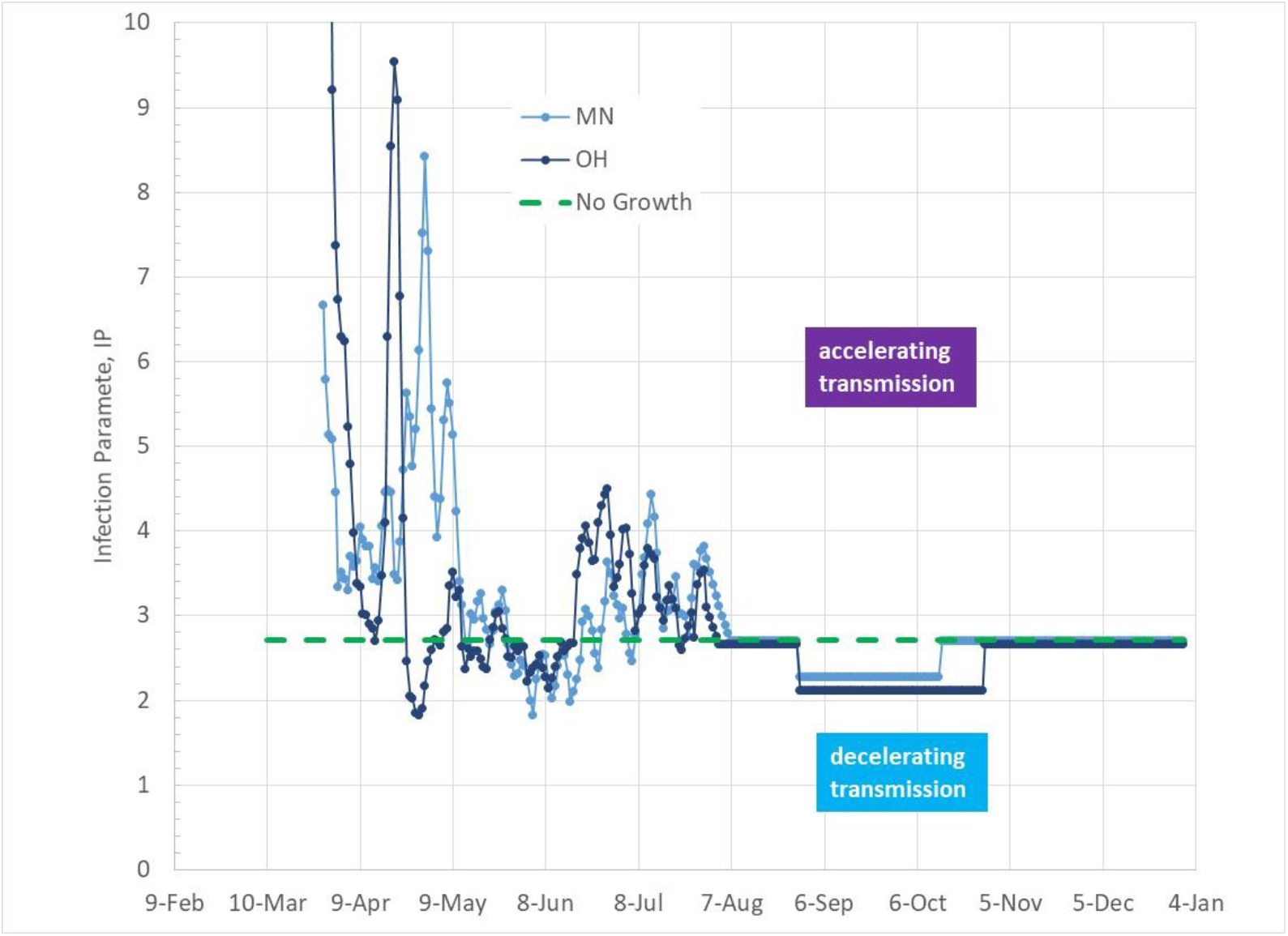
Infection Parameter projections for MN and OH showing impact of outdoor temperature without school re-opening from August through December 2020.

**Figure 16.**
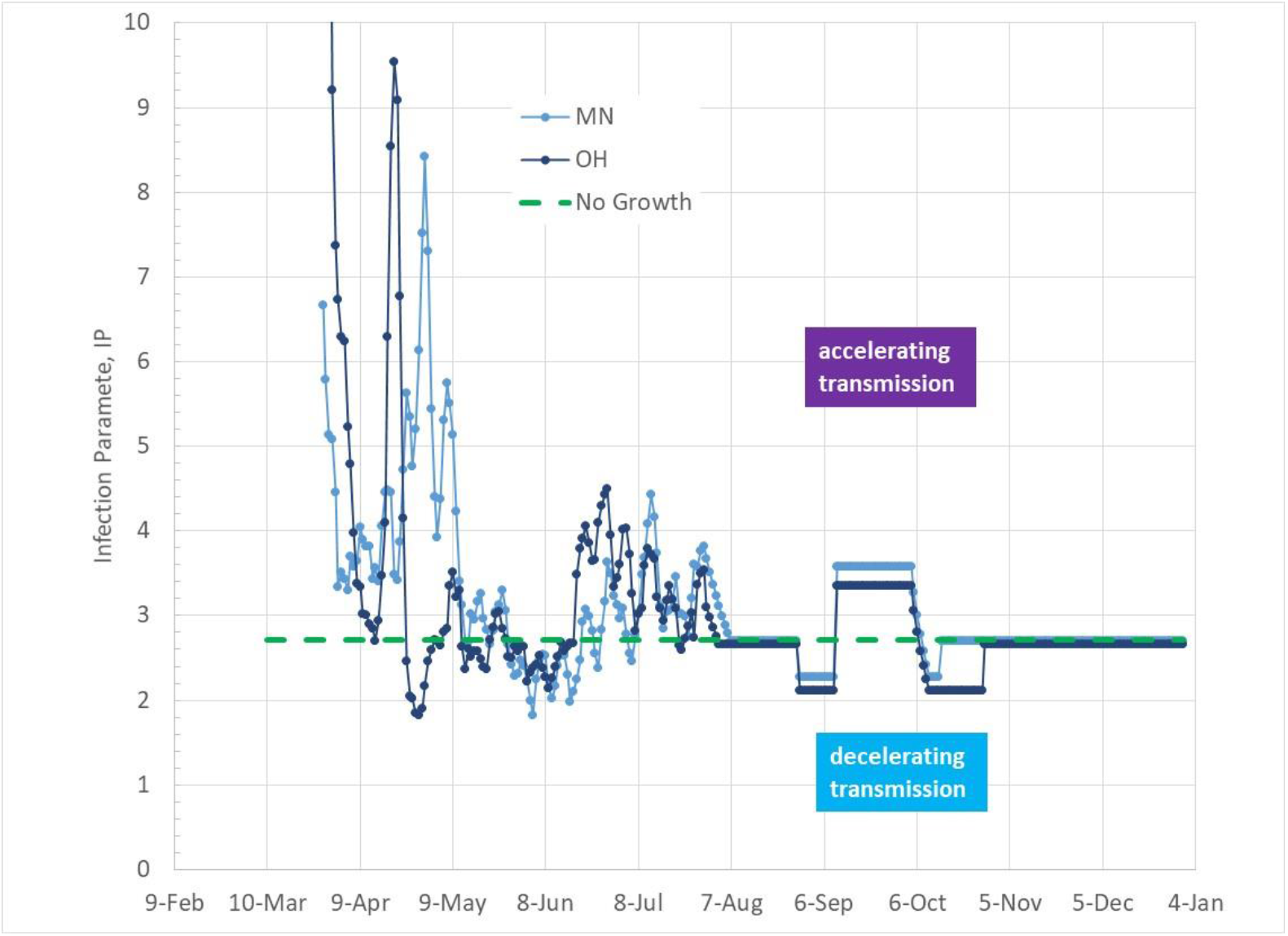
Infection Parameter trends for MN and OH including the effect of outdoor temperature coupled with a 20% SDI reduction during September.

**Figure 17.**
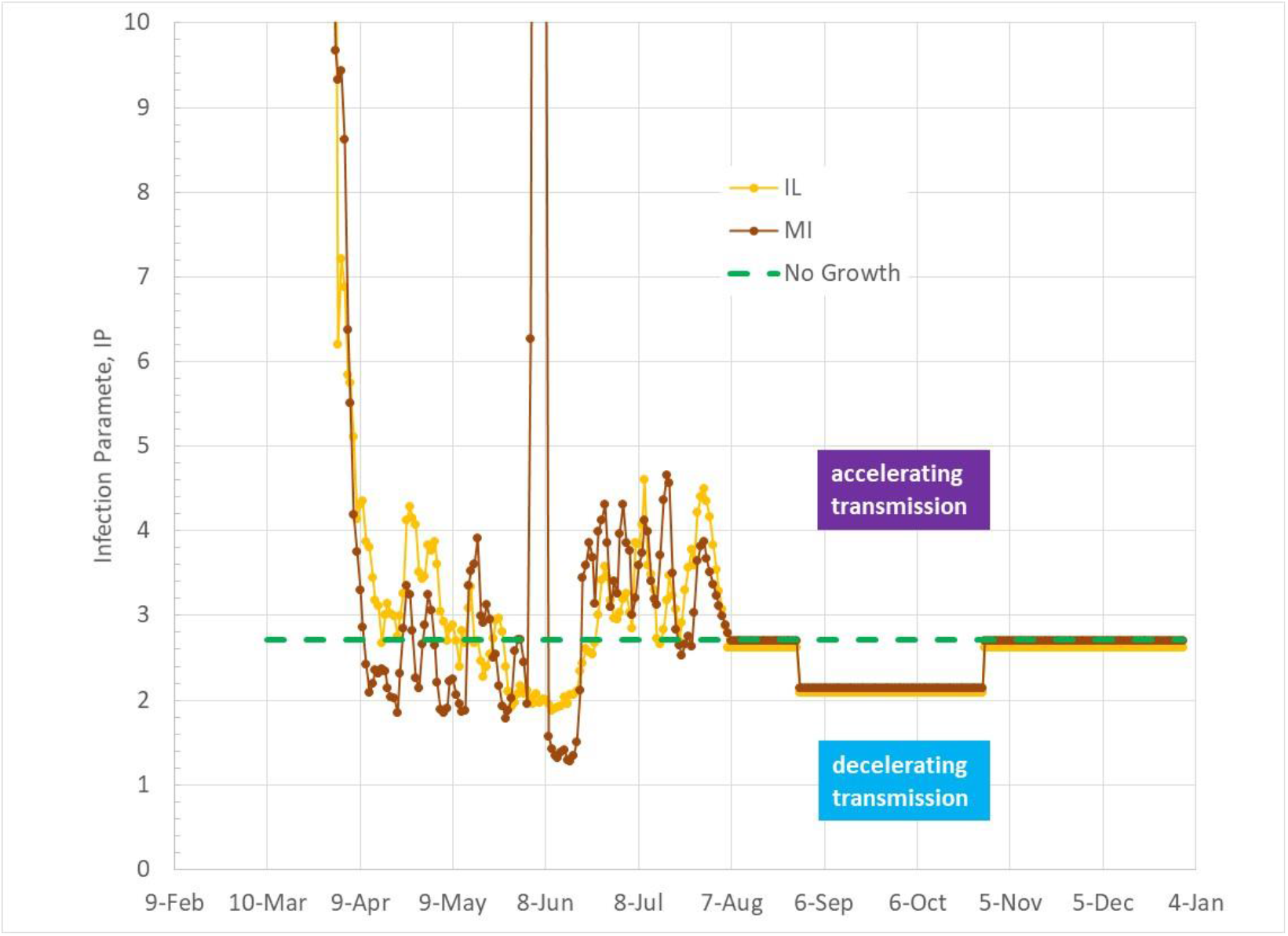
Infection Parameter projections for IL and MI showing impact of outdoor temperature without school re-opening from August through December 2020.

**Figure 18.**
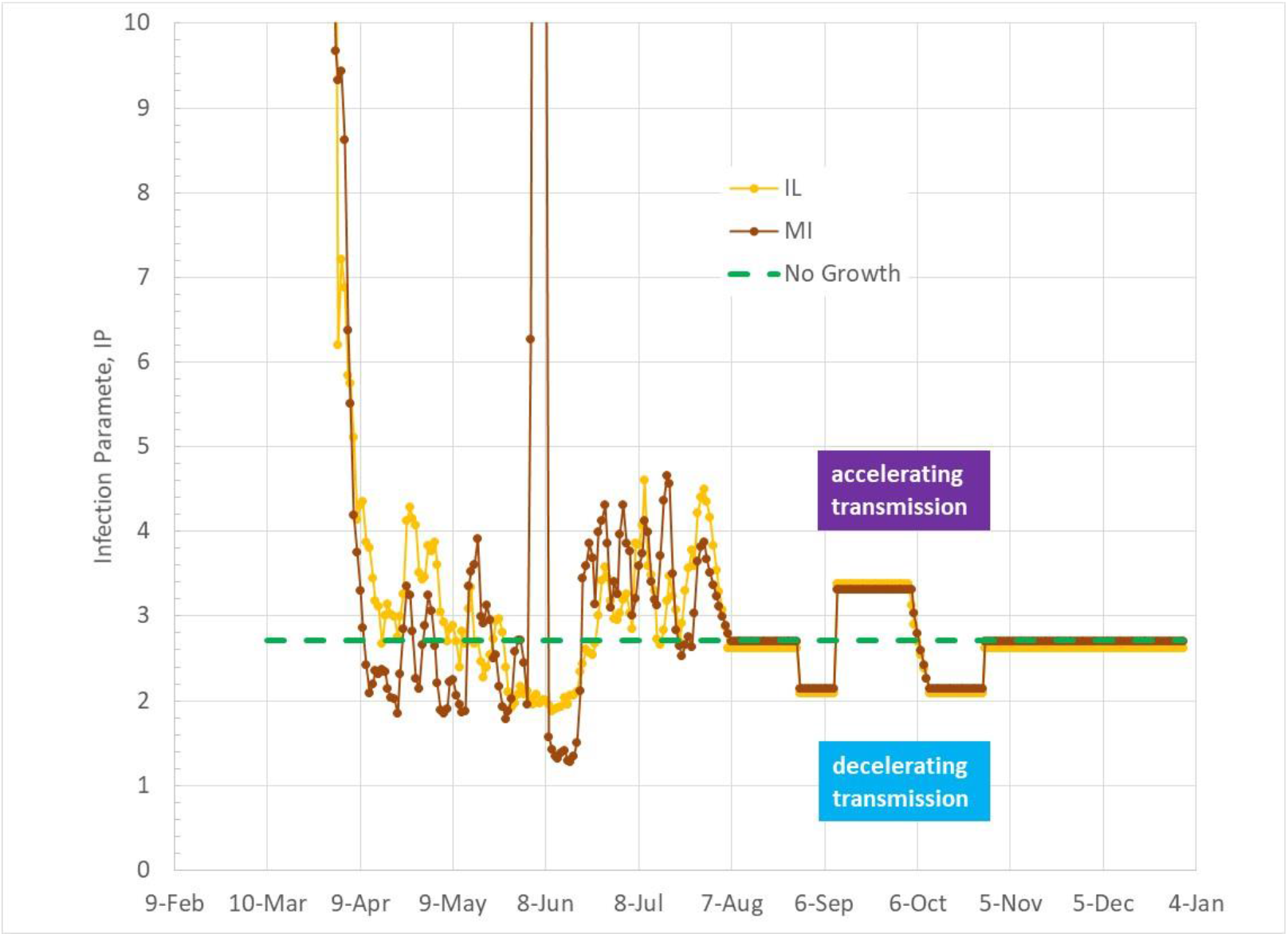
Infection Parameter trends for IL and MI including the effect of outdoor temperature coupled with a 20% SDI reduction during September.

Superimposing the effect of physical school re-opening with an assumed 20% reduction of SDI occurs during NY, WA, and GA swing seasons. The impact of reduced SDI is sufficient to counter the beneficial lowering of transmission efficiency during the swing season, as shown in Figure 12. Note that an early September physical re-opening of schools in GA precedes the outdoor temperature drop in transmission efficiency, resulting in approximately 2 weeks of high increase of IP above the linear infection growth boundary. New York’s September SDI reduction occurs slightly ahead of its beneficial drop in transmission efficiency, while a September reduction of SDI for school re-openings is fully within its fall swing season.

Figures 13 and 14 show IP trends for temperature effect only and temperature effect combined with school re-opening effect for CA, FL and NC. FL and NC do not enter the fall swing season until after September, resulting in high IP levels during September for school re-openings. CA, based on LA weather, enters the swing season within September, tempering the impact of lower SDI caused by school re-openings. NC enters its swing season just after September, and remains in the 50F (10C) to 70F (21C) range until November. FL does not drop below 70F (21C) until November, remaining in the 50F (10C) to 70F (21C) range through the rest of the year.

Figures 15 and 16 show IP trends for OH and MN, and Figures 17 and 18 show IP trends for IL and MI. These midwestern states display similar movement into the fall swing season temperature window in August, and exiting the temperature window in October as temperatures drop below 50F (10C). All four states drop below 70F (21C) about the same time near the end of August. MN drops below 50F (10C) in the beginning of October while IL, MI, and OH drop below 50F (10C) near the end of October. All four states beneficial outdoor temperature drop in transmission efficiency occurs before the assumed September school re-opening, remaining at a lowered transmission efficiency beyond the assumed September reduction of SDI.

We note that the four cases studied combined with the variety of climatic conditions provides examination of SDI re-opening reductions that occur on either side as well as within the expected outdoor temperature swing season.

## Discussion of Results

Table 1 and Figures 19 to 26 show prediction results for the four case combinations of swing season temperature effects and physical school re-opening effects. Overall, physical re-opening of schools increases Covid-19 infections. Two factors are important for reducing the effects of school re-openings. First, for regions with swing seasons that largely surround the assumed September school re-opening period, reduced disease transmission efficiency during the swing season helps mitigate infection growth increases. Second, states that had early infection growth, such as New York and Michigan, with low numbers of current infectious subjects are less prone to accelerated infection growth. In contrast, states such as FL and CA that are currently experiencing high infection growth with large numbers of infectious subjects are much more sensitive to school re-openings.

**Table 1.**
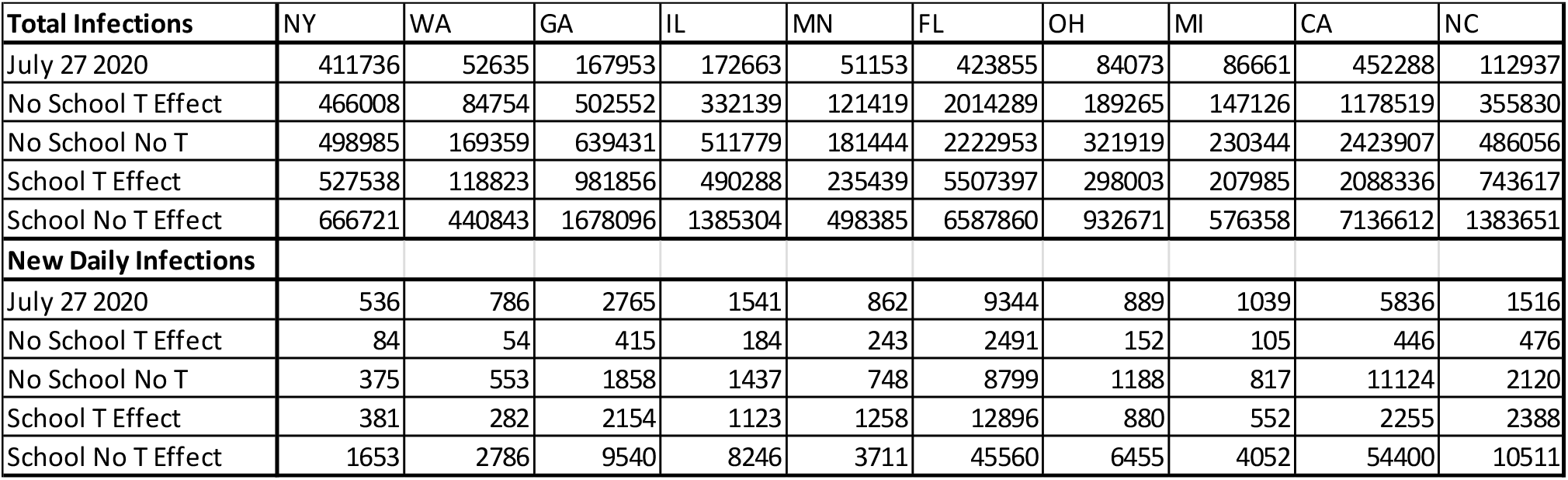
Total Infections and New Daily Infections as of July 27, 2020 and predicted for December 31, 2020 for 10 US States.

**Figure 19.**
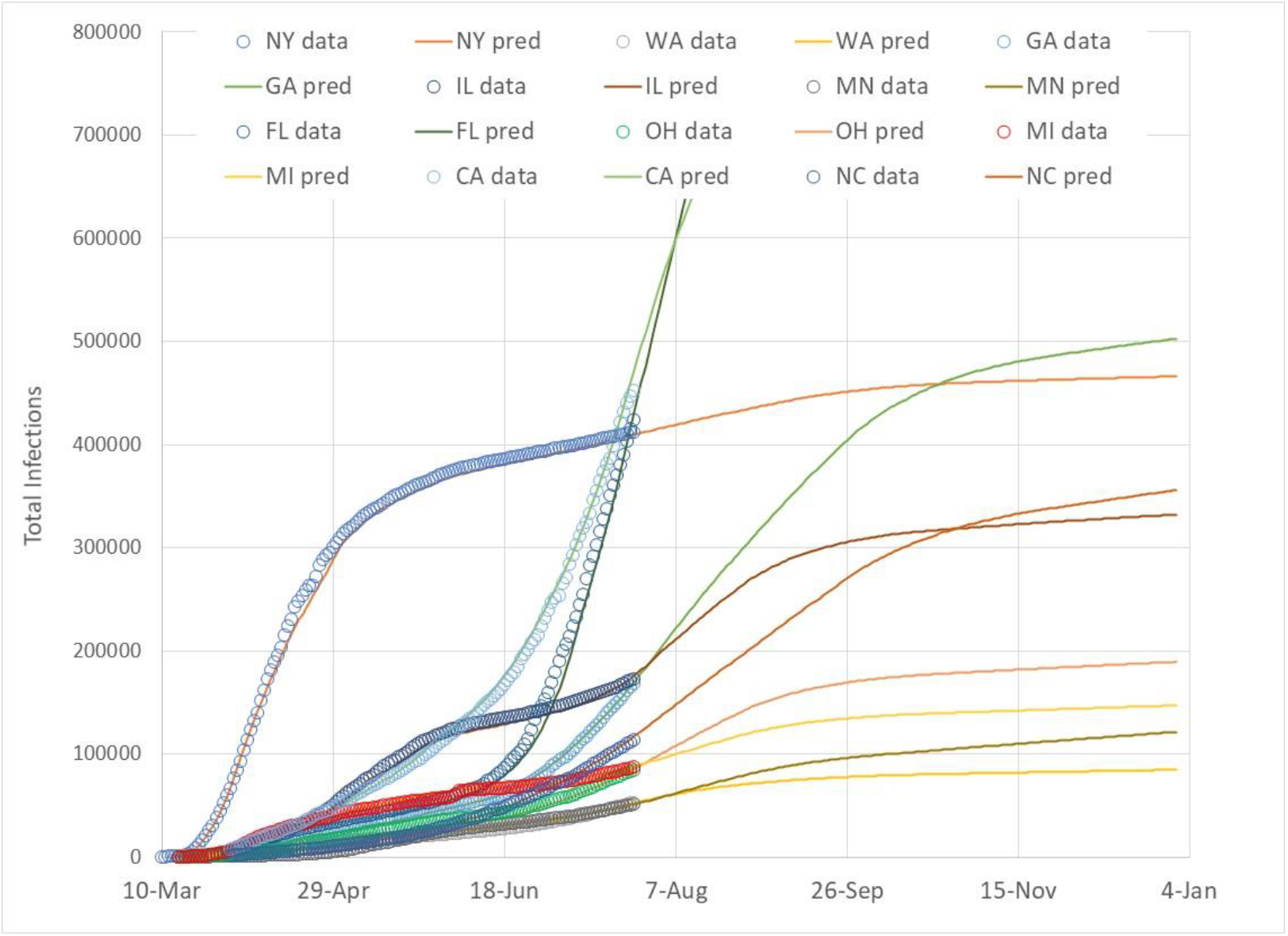
Total infections for 10 US States with actual data (3) and predicted trends from August through December 2020 based on no physical school re-opening, but fall outdoor temperature effect.

**Figure 20.**
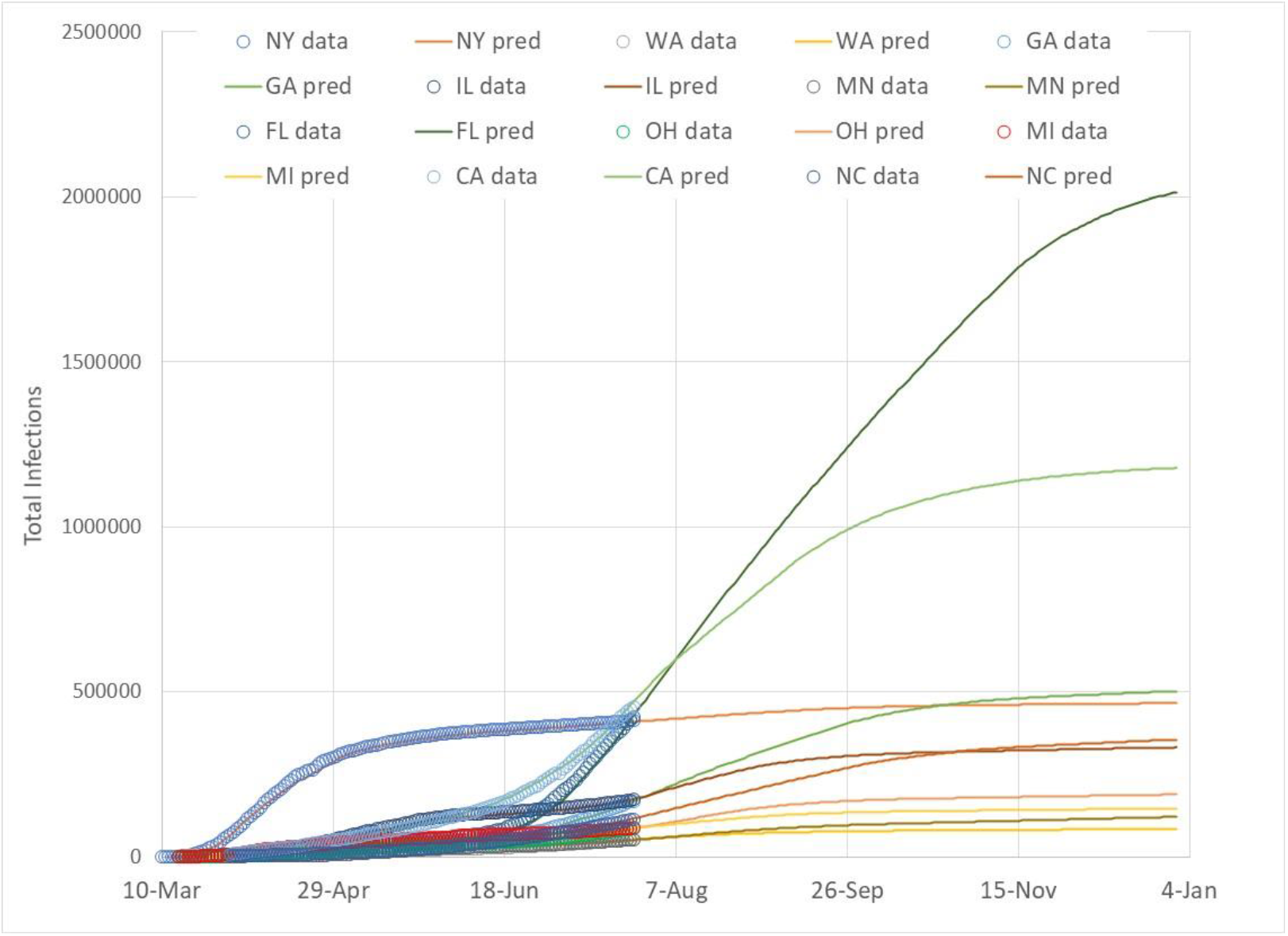
Expanded view of total infections for 10 US States to show potential infection growth paths of CA and FL with actual data (3) and predicted trends from August through December 2020 based on no physical school re-opening, but fall outdoor temperature effect.

**Figure 21.**
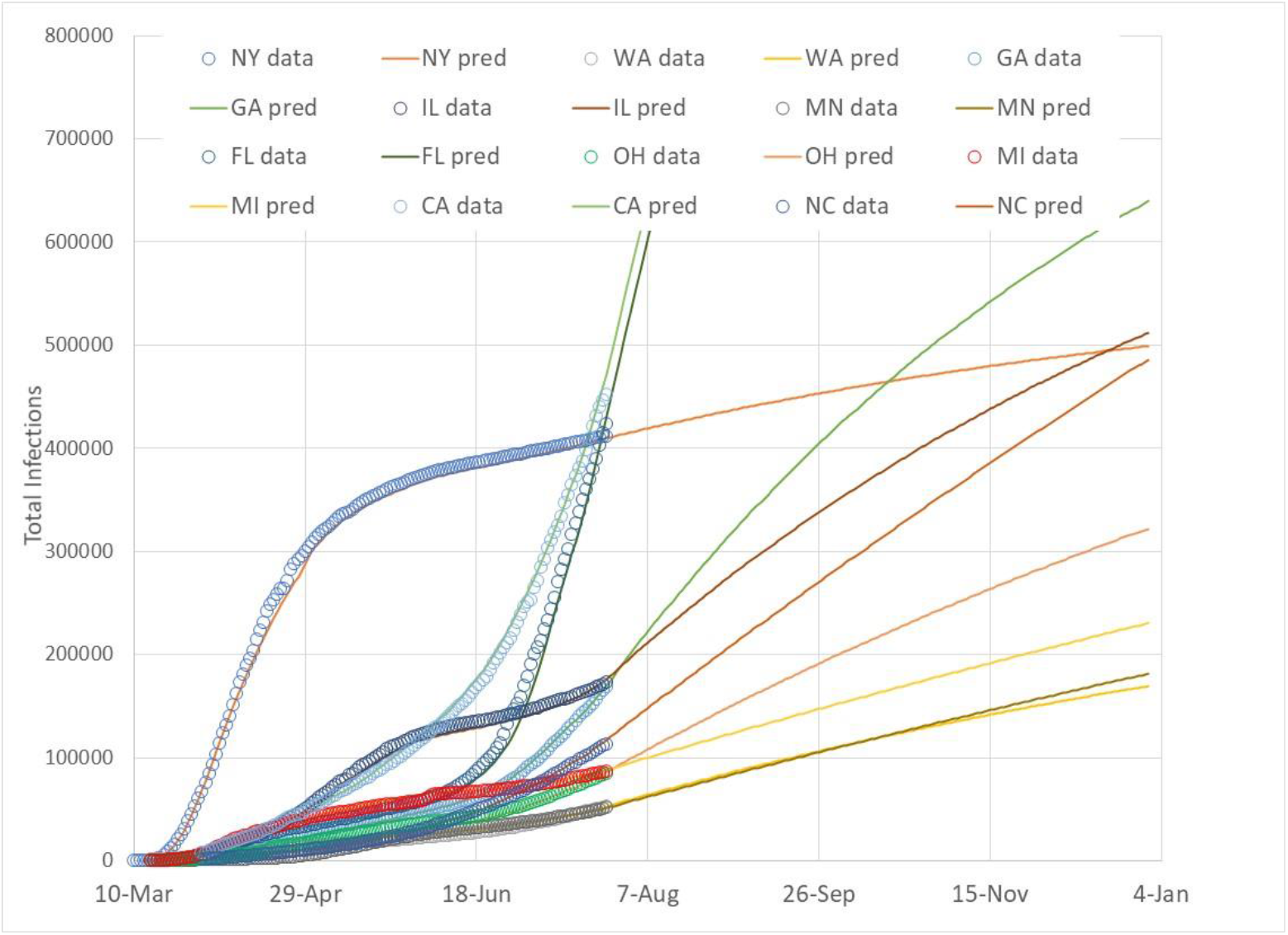
Total infections for 10 US States with actual data (3) and predicted trends from August through December 2020 based on no physical school re-opening, and no fall outdoor temperature effect.

**Figure 22.**
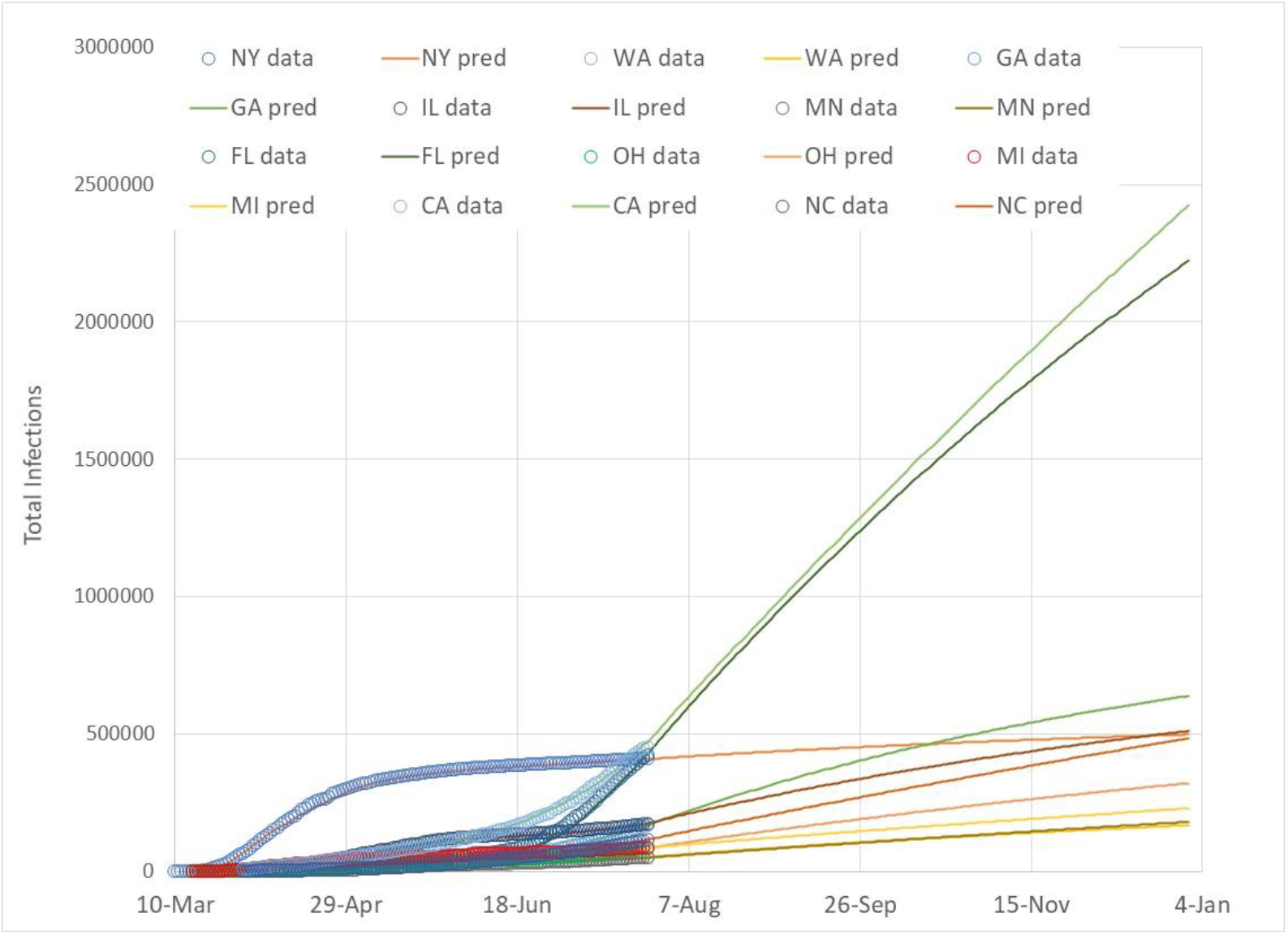
Expanded view of total infections for 10 US States to show potential infection growth paths of CA and FL with actual data (3) and predicted trends from August through December 2020 based on no physical school re-opening, and no fall outdoor temperature effect.

**Figure 23.**
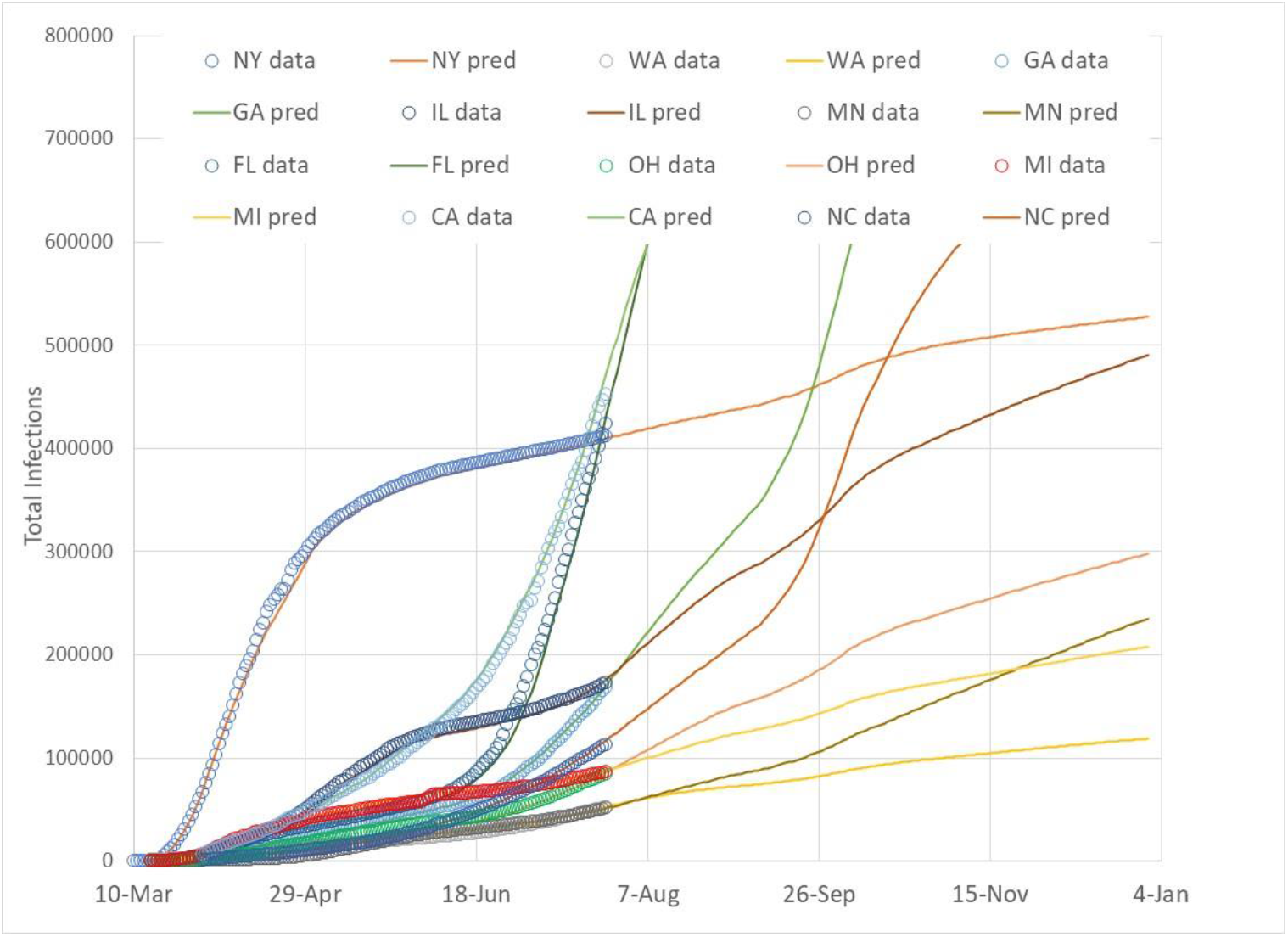
Total infections for 10 US States with actual data (3) and predicted trends from August through December 2020 based on September physical school re-opening with fall outdoor temperature effect.

**Figure 24.**
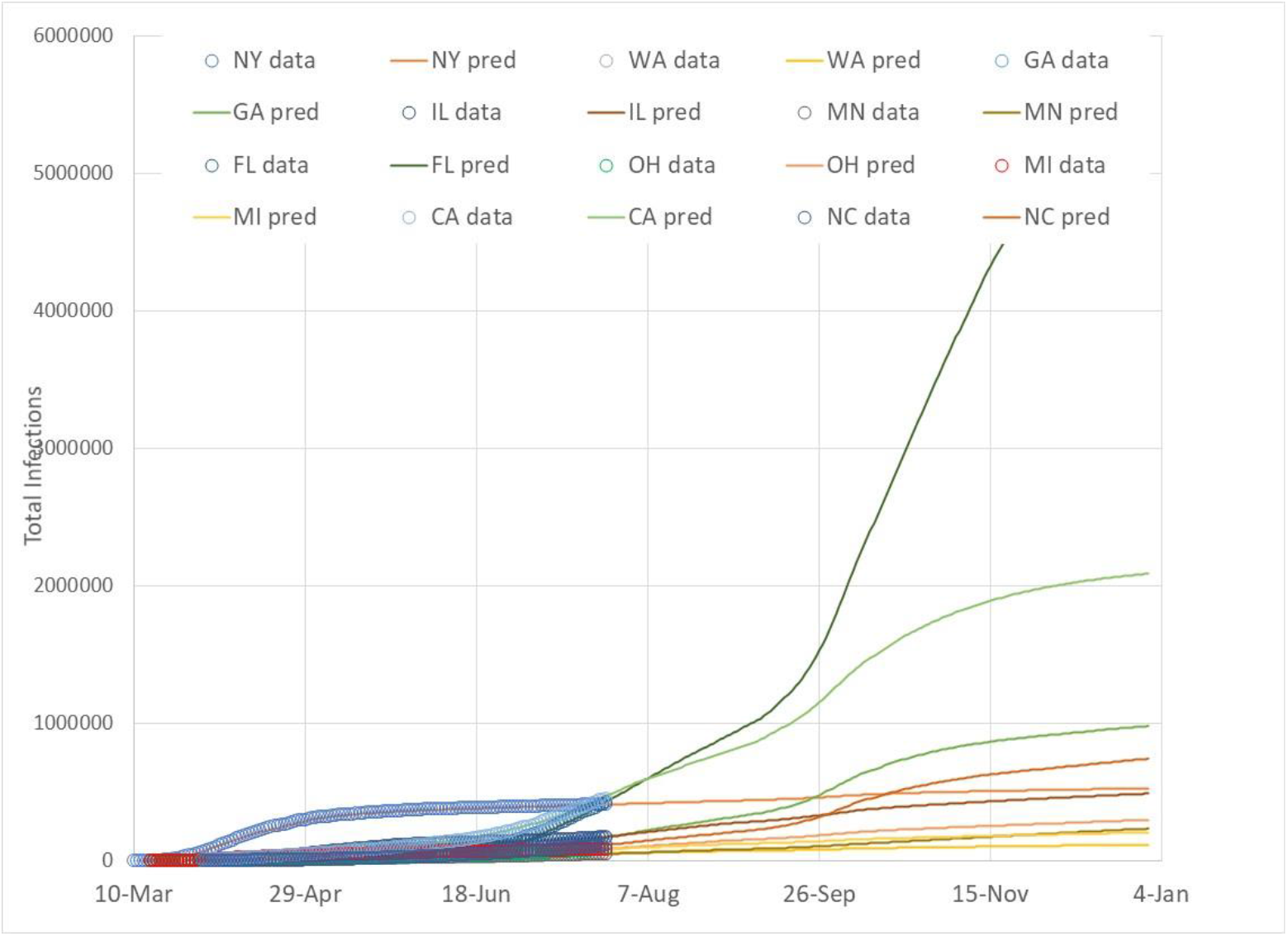
Expanded view of total infections for 10 US States to show potential infection growth paths of CA and FL with actual data (3) and predicted trends from August through December 2020 based on September physical school re-opening with fall outdoor temperature effect.

**Figure 25.**
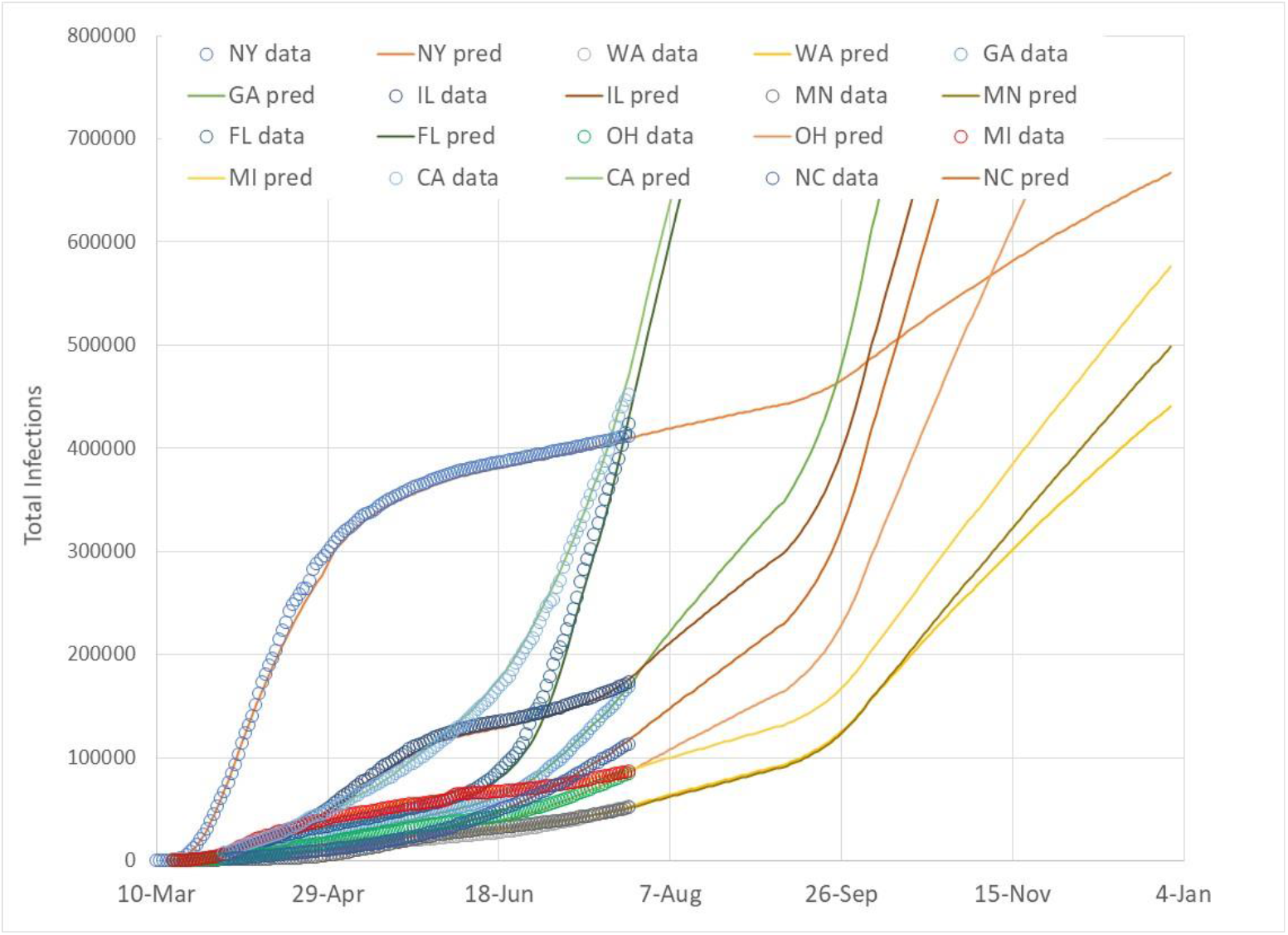
Total infections for 10 US States with actual data (3) and predicted trends from August through December 2020 based on September physical school re-opening with no fall outdoor temperature effect.

**Figure 26.**
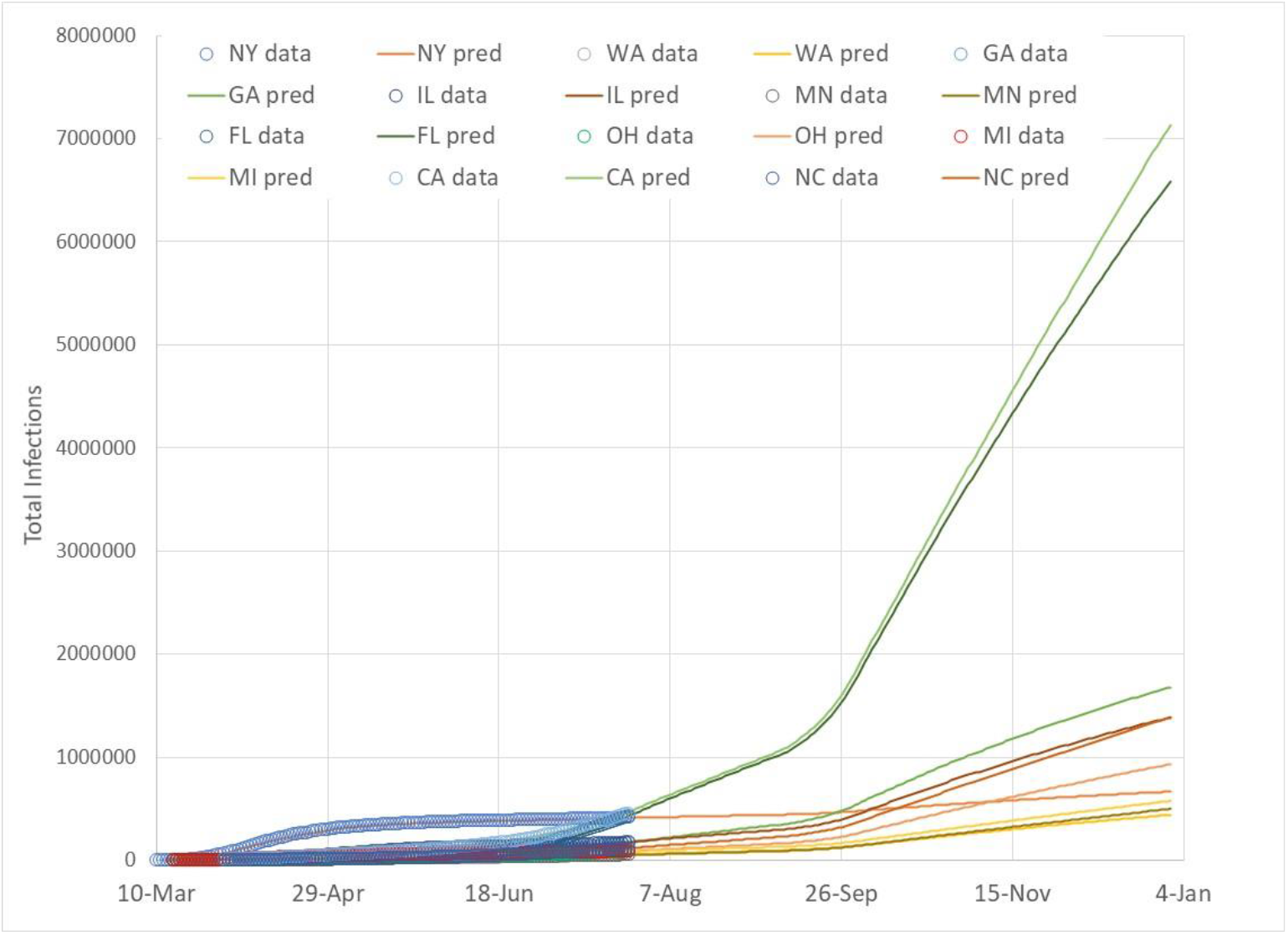
Expanded view of total infections for 10 US States to show potential infection growth paths of CA and FL with actual data (3) and predicted trends from August through December 2020 based on September physical school re-opening with no fall outdoor temperature effect.

NY, CA, and FL are interesting to discuss together because these high population states have total infections in the 400,000 to 500,000 as of the end of July 2020. NY experienced rapid, uncontrolled infection growth in the spring, and has since been able to keep new infections low, while FL and CA are currently in the midst of rapid infection growth. New York has much fewer infectious persons than either California or Florida because its early infectious cases are no longer infectious. Infection growth rates are proportional to an infection growth constant (derived from the Infection Parameter, (1)) times the number of infectious persons. Therefore, an increase of IP above the linear infection growth boundary (IP=2.72) more significantly impacts states with higher numbers of currently infectious persons.

Table 1 summarizes current total infection numbers and current daily new infection cases as of the end of July 2020, and predicted total infection cases and daily new infection cases at the end of December 2020. NY is projected to grow by somewhat more than 10% in total infections by the end of December without school re-openings, but including a beneficial impact of fall swing season temperatures in comparison to a 20% total infection growth for NY without school re-openings and no swing season temperature effect. Daily new infections, without school re-openings for cases with and without swing season temperature effects, would decrease from current new infections based on the assumption of a steady IP value slightly below the linear growth boundary.

Physical re-opening of schools in September with an assumed SDI reduction of 20% may result in 30% (beneficial swing season temperature effect) to 60% (no swing season temperature effect) for NY relative to end of July total infection cases as listed in Table 1. Note that the impact of September school re-openings for all other states investigated more than double relative to end of July total infections, with some states, such as CA, FL, OH and NC increasing by more than 10 times if no beneficial swing season temperature effect occurs.

As infections grow through the fall, US citizens in states with large predicted infection numbers, such as CA and FL, may choose to deviate from the assumed linear growth path by implementing measures such as mandatory mask wearing and increased testing, tracking and isolating that moves the populace to a sustained IP level below 2.72, resulting in infection decay. Only time will tell. This report will be updated in early September to re-assess the trends observed for each state.

Figures 19 and 20 show predictions for no school re-openings with a swing season temperature effect. Figure 20 is an increased scale to show FL and CA growth relative to the other 8 states. Note that total infections modestly grow for most states other than FL and CA because warmer states have delayed outdoor temperature drops into the swing season range. FL and CA continue linear infection growth trends until their outdoor temperatures drop below 70F (21C). FL experiences the longest period of linear growth because it is the last region to reach 70F (21C). Note that all states follow a linear growth path that is an extension of their late July infection growth path until they reach a time when their outdoor temperature drops below 70F (21C). Infection growth rates then drop with a decay in daily infection cases due to a reduction below the linear growth limit. Once the swing season ends, each state then resumes its linear growth path, *but at a lower slope than before*. The linear infection growth boundary is not dependent on infection slope (infection growth rate) magnitude. Note that FL is the only state assumed to have a swing season that continues to the end of December.

The observed tendency of the US could result in relaxed infection control measures during the beneficial swing season decrease of disease transmission efficiency. That is, if a populace observes decreasing numbers of daily infection cases during the swing season, and adopts less stringent control measures, IP could return to 2.72 or greater, resulting in accelerated infection growth once the swing season ends.

High predicted infection levels for FL and CA, ranging up to 6 to 7 million infections for the case with school re-opening and no beneficial swing season temperature effect represents less than 50% of those state populations. Infection equilibrium (“blanket immunity”) for Covid-19 without vaccinations is estimated to be as high as 70% of a population group. That is, the worse case scenario for these predictions continues to be in the early stage of disease progression.

Figures 21 and 22 show total disease infection paths for the case of no school re-opening and no swing season temperature effect. The assumed tendency of the US populace to oscillate across the linear infection growth path results in a linear infection growth to the end of December. All states, based on the slope of their current infection paths, continue along that line of growth. The previous case that includes a beneficial swing season temperature effect lowers total infections by a significant amount. Figure 22 is an expanded view of Figure 21 showing CA and FL infection growth to the end of December 2020.

Figures 23 and 24 show infection trends for the case with September school re-openings coupled with a fall swing season temperature effect. Infection path curvatures are most complex for this case as school re-openings and beneficial swing season temperatures move the Infection Parameter in opposite directions. Depending on a location’s climate, infection path slopes may either increase initially if school re-openings occur before swing season temperatures, or decrease if swing season temperatures occur before September school re-openings. The final slope of the infection growth path will be determined by infection path conditions as both school re-opening and swing season temperature effects are passed. Beyond that time, the assumed linear growth tendency of the US occurs.

Figures 25 and 26 present results for the final case with school re-opening and no swing season temperature effect. Similar, but opposite in trends to the first case with no school re-opening with a swing season temperature effect, each state follows a linear infection growth path until September when the SDI is reduced 20% as schools re-open. Each state follows a linear infection growth path through August, similar that shown in Figure 1 for the US as a whole. The reduced SDI representing school re-opening increases IP above 2.72, causes accelerated infection growths through September. After September, IP returns to 2.72, continuing on a new linear based on the final infection slope at the end of September. This case represents the worse of all cases because the accelerated infection growth does not have a swing season beneficial effect to counter the IP increase.

## Summary

Overall, the four cases for 10 US States demonstrate the sensitivity of infection growth through the end of 2020. Infection predictions that range from 1.1M to 7.1M, as is the case for California, is not reassuring, however, this range of infections is not due to the prediction model accuracy but rather to the uncertainty of human behavior.

Will the US continue along a linear infection growth path that is characteristic of a populace that lacks a coherent, coordinated infection control policy? Linear growth paths represent an oscillatory behavior of a populace that alternately switches from improved infection control (increased SDI and lower G) when accelerated infection growth reports alarm citizens, to a more relaxed state (decreased SDI and higher G) as pressure to re-open businesses, schools and public gathering places builds.

The unusual, real time nature of the ongoing pandemic coupled with technology that allows timely posting of non-peered review reports has not happened before. In this regard, this report predicts infection trends based on observations that may prove to be true or false. Monthly updates to this report will be provided to compare future data with the predicted infection path data.

## Data Availability

All data used for the report are publicly available.

https://www.worldometers.info/coronavirus/

http://91-divoc.com/

https://data.covid.umd.edu

